# Identification of Patterns in Epidemic Cycles and Methods for Estimating Their Duration: COVID-19 Case Study^1^

**DOI:** 10.1101/2020.07.13.20153080

**Authors:** Eduardo Atem de Carvalho, Rogerio Atem de Carvalho

**Affiliations:** Advanced Materials Laboratory, Universidade Estadual do Norte Fluminense, Brazil; Innovation Hub, Instituto Federal Fluminense, Brazil

## Abstract

This paper presents several epidemic cycles of COVID-19 that have practically ended in countries, states and cities and normalize them through simple and well-known numerical methods. It is evident that there is a practically universal pattern between them, in a triangular shape. It is also possible to find similar cycles with very close scales and thus use cases with cycles already closed to predict the end of the cycles still in progress. Three methods are presented and discussed and the case of the city of Rio de Janeiro, Brazil, is presented in more detail.

## 1) Introduction

With the recent COVID-19 Pandemic, an issue that immediately arose was the length of its cycle. As the virus does not spread instantly across the globe, there will naturally be countries where it will be at a more advanced stage, because it started earlier, and others where it will be more “delayed”. Therefore, the most common way of trying to make this type of prediction is to use the contamination cycle of the places where this contamination started earlier to try to predict how the disease will evolve where it started later. But, in order to make such a comparison, knowing that the sizes of the populations are different, some measures are necessary so that the pattern of the local behavior of the pandemic is properly identified.

Therefore, this paper proposes to use the combination of the use of normalization of epidemic cycles already closed superimposed on values from public databases to reach some conclusions and estimates on the cycle durations for those that are still in progress. The entire theoretical and numerical basis employed is presented, demonstrated and discussed before any projection is made. In this study, the term “epidemic” will refer to a local episode, be it in a city or even in a country. When the term used is “pandemic”, the authors refer to the worldwide phenomenon.

In the beginning of the 21st century, several transnational entities and government agencies have become convinced of the inevitability and have been preparing for a devastating pandemic along the lines of the medieval Black Death [1]. The reason for this is beyond the scope of this work. Also, and practically inevitably, along with the discussions comes the predictions, many of them dire [2], and based on supposedly almost infallible mathematical models. For it was enough to have an episode of greater magnitude, such as COVID-19, for all predictions based on these models to fail [3], but the panic generated and the measures that followed will leave consequences for generations.

Based on the observation of available data and without attempting to elaborate any model of a deterministic predictive nature, this work analyzes several cases of countries that have already faced what at the moment appears to be the critical cycle of the disease, allowing some more obvious patterns to be detected. Discussions of the aggravating or mitigating factors of the disease also escape the modest scope of this work.

Finally, this paper will only work with the number of registered deaths. The actual number of non-fatal COVID-19 cases, in their various intensities, is impossible to determine in a correct and absolute way, since this would imply that all populations would be tested continuously, or at least continuous samples and significant changes were tested over the duration of local epidemics, which was not done [4], although some countries, such as Germany, tested significant amounts of their population at the beginning of the pandemic. Thus, the number of deaths, although not perfect, is less subject to factors such as those described above.

## 2) On the Nature of the Observed Data

The attempt to describe the different epidemic cycles that make up the current pandemic often comes up against the quality of the data that is made public. Deaths occur on different dates than those recorded daily by different governments and health authorities. This is inevitable at first and will be corrected over time, so that the correct dates when the deaths occurred will become available. At the present time this is an ongoing process. To introduce and comment on and present the known methodology previously cited as the Moving Average Method, the case of Sweden is presented, which serves as an example, since the first data available up to the end of June 2020, brought the deaths on the days of registration, in this way, the distribution of fatalities suffers a distortion that generates a “saw” appearance in the graphs of the values presented (Figure 1), and on weekends there is a clear absence of deaths, followed by an explosion of those values at the beginning of the weeks. One way around this effect is to apply the so-called Moving Average Method (MAM), in which the daily value of deaths is replaced by the average of the previous 6 days plus of that date. With the disclosure by Sweden of the correct dates of death, one can compare the effectiveness of MAM with the actual values.

**Figure 1.**
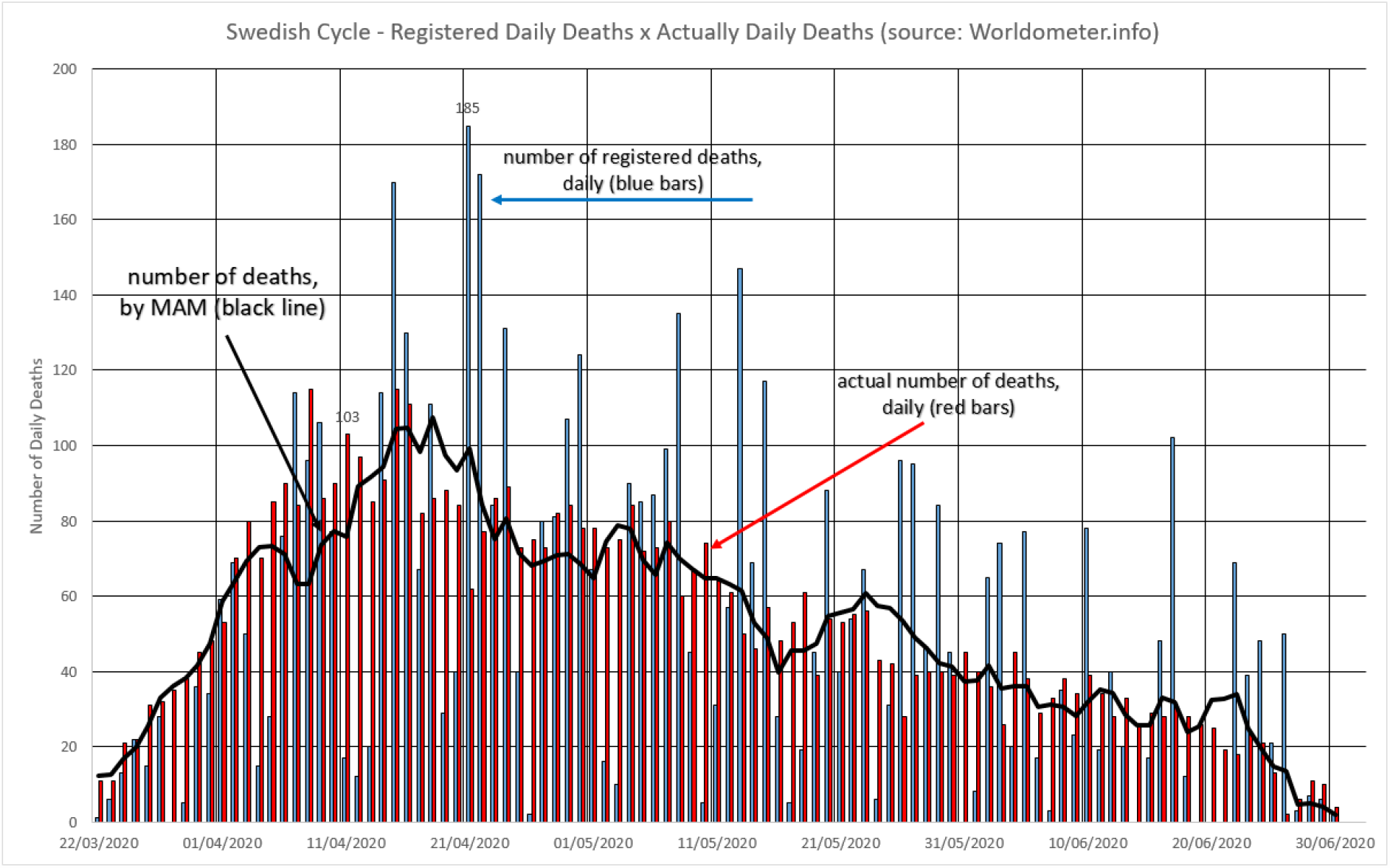
Graphical match between data shown before for Sweden (number of registered deaths, daily – blue bars [6]) and the updated for actual number of daily deaths (red bars [7]). Black line represents the MAM figures.

In order to judge whether MAM is an applicable method to minimize the effects of seasonality on the dates of registration of deaths, the statistical criterion known as R^2^ or R-Square [5] will be used, which in this case determines how well the MAM values describe the true data, that is, if MAM is used, how close we will be to the distribution of deaths according to the actual date that occurred. The value of R^2^ can vary between 0 and 1 and in general, the closer to 1 the value of R ^2^ is, the better the performance of MAM. R^2^ is defined as follows:

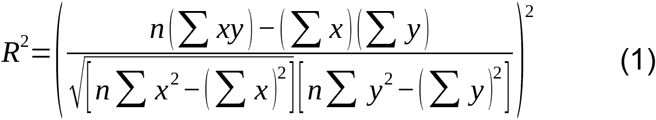

Where, in this case, x are the points containing data referring to the actual number of deaths that occurred on each date of the epidemic cycle and y are the values generated by the Moving Average Method and attributed to the observed days. Three basic initial models were tested for the Swedish case:

a. Moving Average having the central position as the defined value (the one shown in the graphs). For example: in a 7-day week, starting on Sunday and ending on Saturday, the average of the 7 values will be computed in the Wednesday value. This is the case of the Moving Average Method with Central Value (MAMC).
b. Moving Average with the initial position as the defined value. In other words: in a 7-day week, starting on Sunday and ending on Saturday, the average of the 7 values will be computed in the Sunday value. This is the case with the Moving Average Method with Initial Value (MAMI).
c. Moving Average with final position as the defined value, in other words: in a 7-day week, starting on Sunday and ending on Saturday, the average of the 7 values will be computed in the Saturday value. This is the case of the Moving Average Method with Final Value (MAMF).

Table 1 shows the values obtained for the cases a, b and c described above and compares them.

**Table 1.** Basic statistical variables for the number of deaths in Sweden in the period from 03/22/2020 to 06/30/2020.

| | Average | Sample Variance | $R^2$ | Obs. |
| --- | --- | --- | --- | --- |
| Deaths Real Date | 53 | 758 | 1,0000 | Reference Values [7] |
| Deaths Record Date | 52 | 1974 | 0,2409 | Initially available values [6] |
| MAMC | 52 | 626 | 0,7241 | Calculated according to (a) |
| MAMI | 52 | 648 | 0,8448 | Calculated according to (b) |
| MAMF | 52 | 645 | 0,5472 | Calculated according to (c) |

Obs.: Decimal places are necessary because the value of $R^2$ is often very close to the unit and its change can only be perceived when operating with a minimum number of decimal places. Values outside this period were excluded from the calculations.

It is clear that the Initial Moving Average Method (described in item (b) above) is the one that best describes the studied phenomenon, as would be expected, since deaths can only be registered after they have happened, creating an inevitable delay in the values disclosed during an epidemic cycle. However, it is also observable that the proposed method (MAMI) manages to convert the registration values in an acceptable way to values close to the true ones. This study will use MAMI in the graphs and simulations below.

## 3) The Chinese Epidemic Cycle

Early in the pandemic, its point of origin, China, completed the epidemic cycle in 8 weeks and data on infected and fatalities became available for analysis [8]. The distribution of fatalities per day of the epidemic seemed to follow the classic Gaussian curve and the duration of the cycle indicates that the disease followed its cycle as predicted (Figure 2) and expected by several models and sources with analytical credibility [9]. These facts contributed to several initial catastrophic or overly optimistic predictions and analyzes. Over time, even the predictions of more experienced groups contained margins of error and expectations that reached 300% variation [4]. The discussion on the Chinese cycle is out of the scope of this work, but it shows why some reactions were extreme, either by underestimating the epidemic - by seeing a country with 1.4 billion inhabitants having relatively few deaths, or by predicting apocalyptic growth, by applying the exponential in the rise of the pandemic in the West.

**Figure 2.**
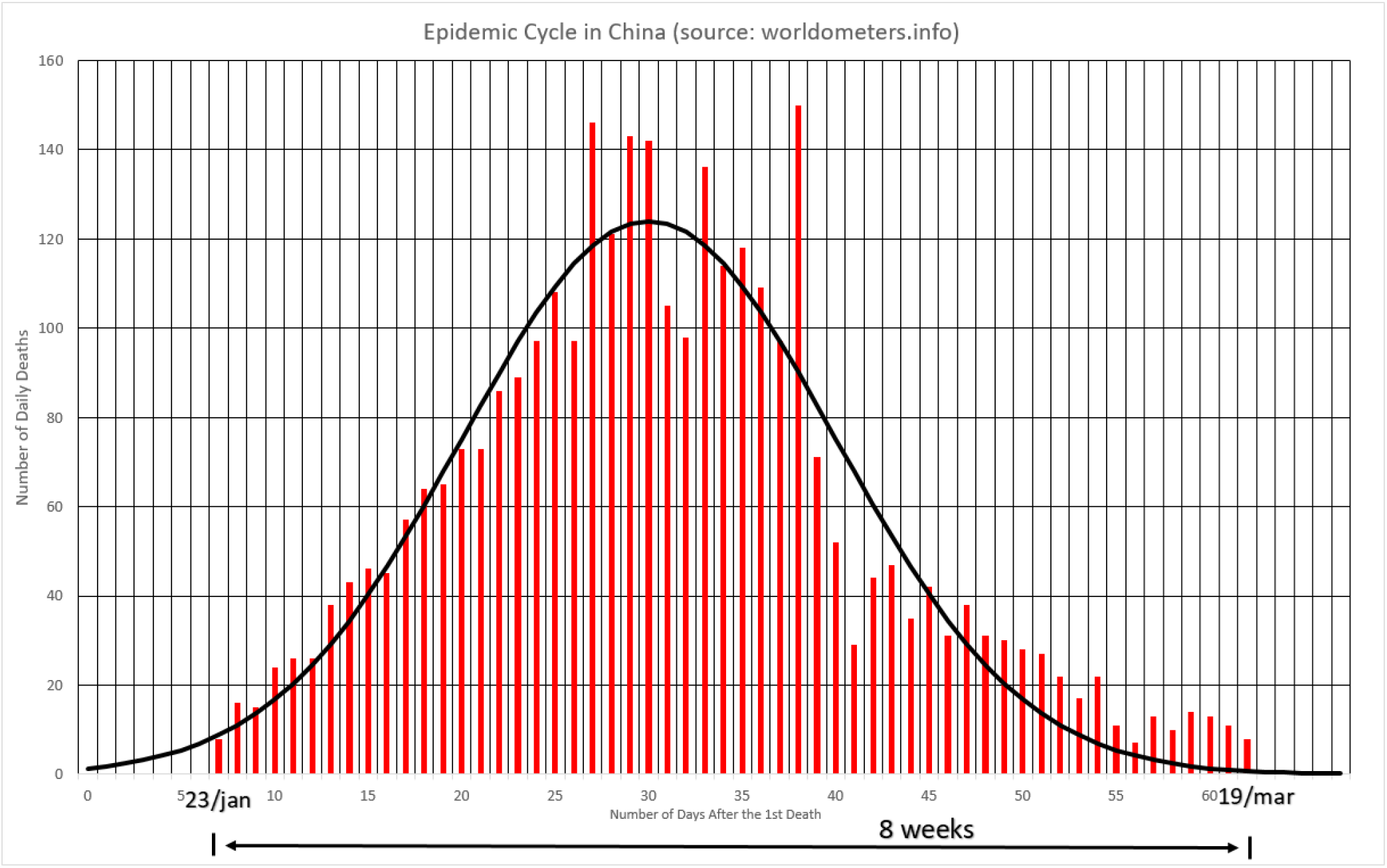
COVID-19 deaths in China.

## 4) Observed Epidemic Cycles

At the time of the writing of this paper (July 2020), several cities, regions, states and countries have already completed what will be defined here as the Most Lethal Cycle of the Epidemic (MLCE), which is when the number of deaths increases daily, on average, until it reaches a peak and then begins to decrease continuously until it reaches a minimum value. After this period, the occurrence of deaths continues intermittently, but relatively small and oscillating, decreasing to certain levels of daily deaths, where it then becomes apparently chronic and presents relatively low, but not zero, values. It is not yet possible to determine whether they are universal phenomena or associated with specific conditions for the evolution of the disease, such as climate, eating habits, genetics, history of vaccination, etc.

### 4.1) Spain

The first large European country to tackle its epidemic outbreak, presented the following cycle during the MLCE, as described in Figure 3. The end of the cycle is considered the last day before the first day with zero deaths [10].

**Figure 3.**
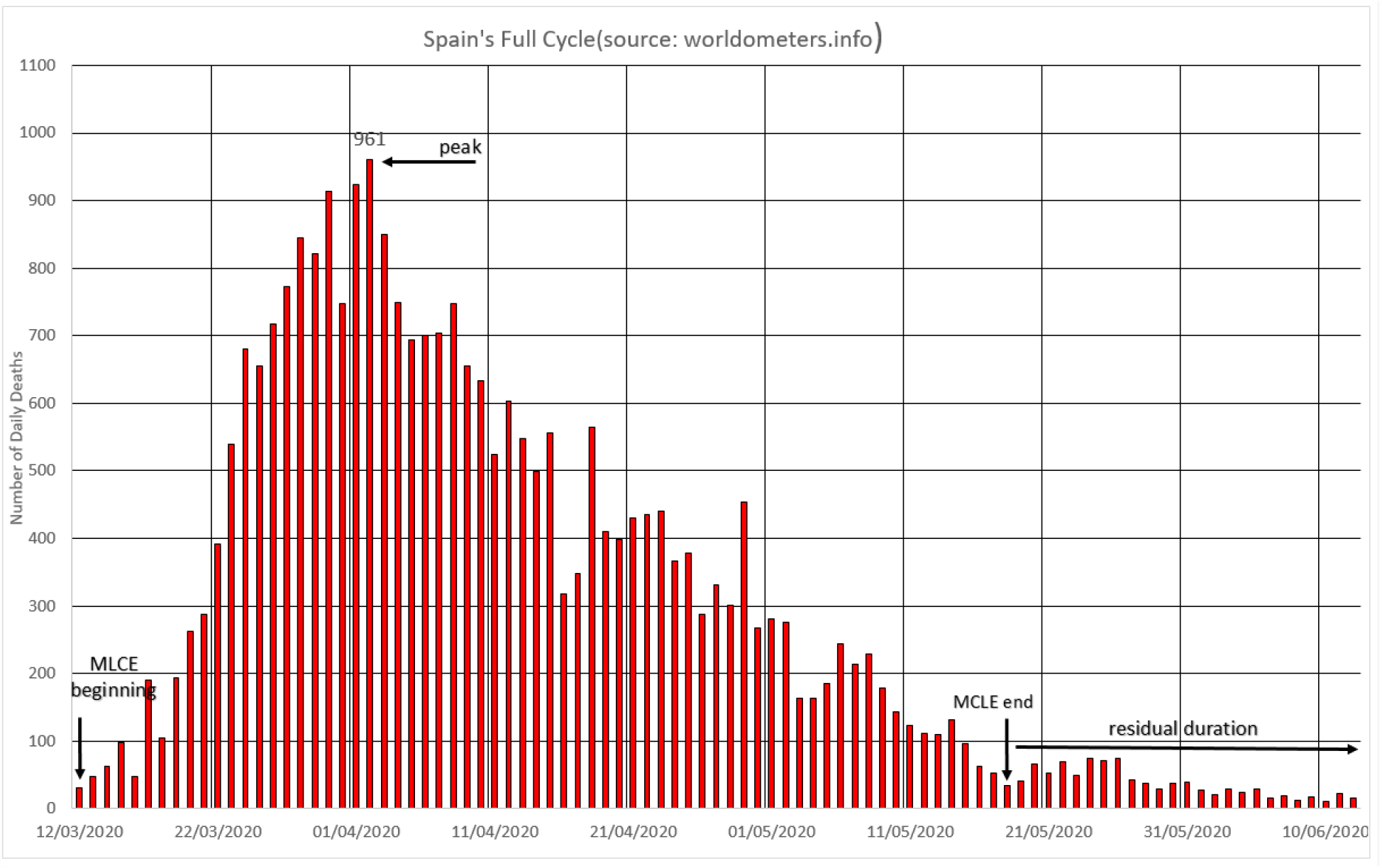
Spain’s MLCE.

### 4.2) The Netherlands

The Netherlands managed to contain the epidemic within its territory and by the time of updating these data (July 3, 2020) it had already reduced the number of fatalities to the units. Figure 4 shows data for the Netherlands [11].

**Figure 4.**
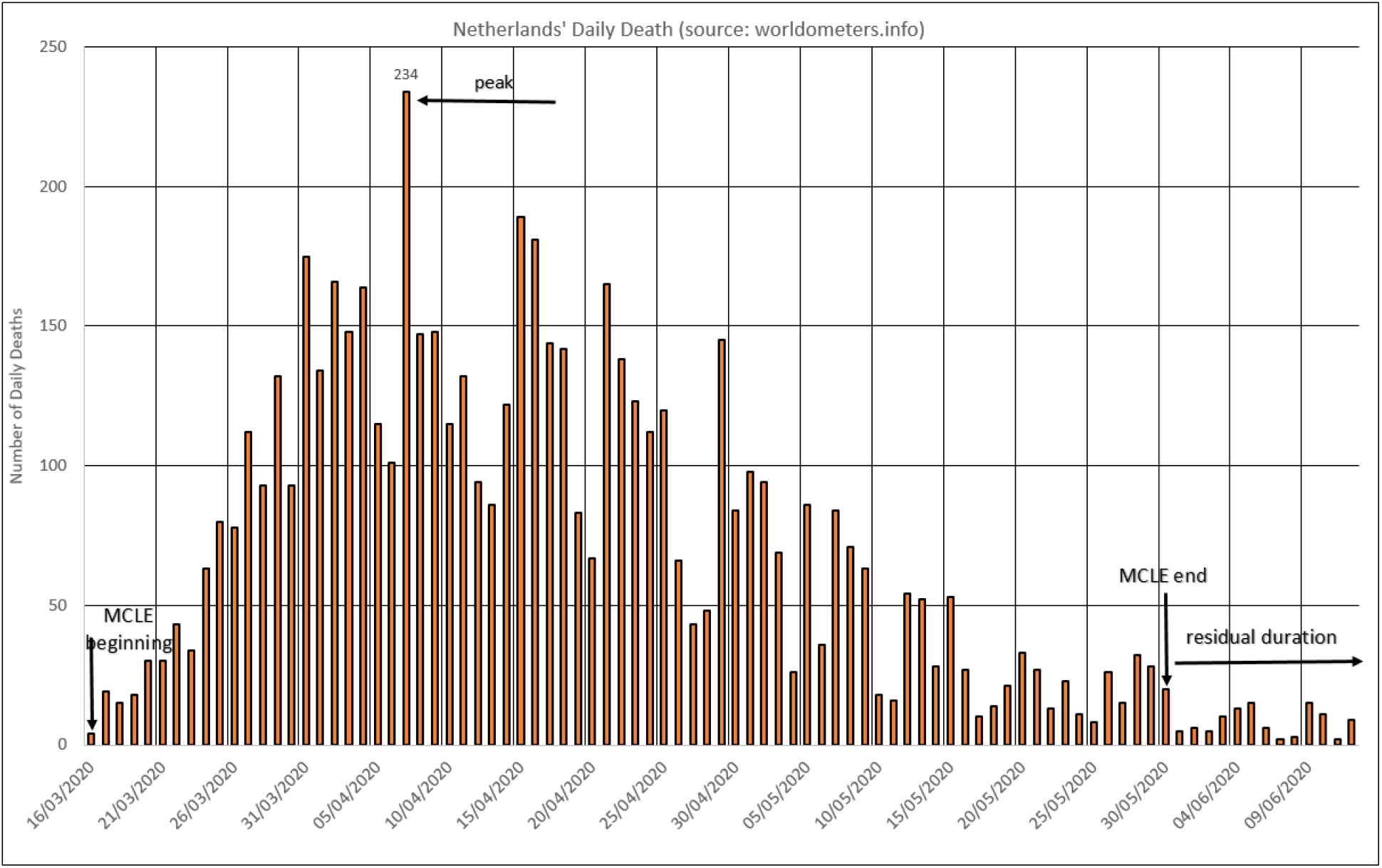
Most Lethal Cycle of the Epidemic – The Netherlands (MLCE).

### 4.3) Italy

A country that was at the European epicenter of the crisis, it has an evolution in the number of deaths (Figure 5) that indicates the overcoming of the MLCE [12]. This work points to the existence of the so-called False Peaks. These are local maximums that were recorded during the cycle of rising or falling in the trend of deaths, but they are not inflection points. In order for a point to be considered as a Peak, it is necessary to register a tendency of a dropping in the number of deaths. This fall will not be linear, but there is an obvious, numerical, and visual trend that indicates such a thing.

**Figure 5.**
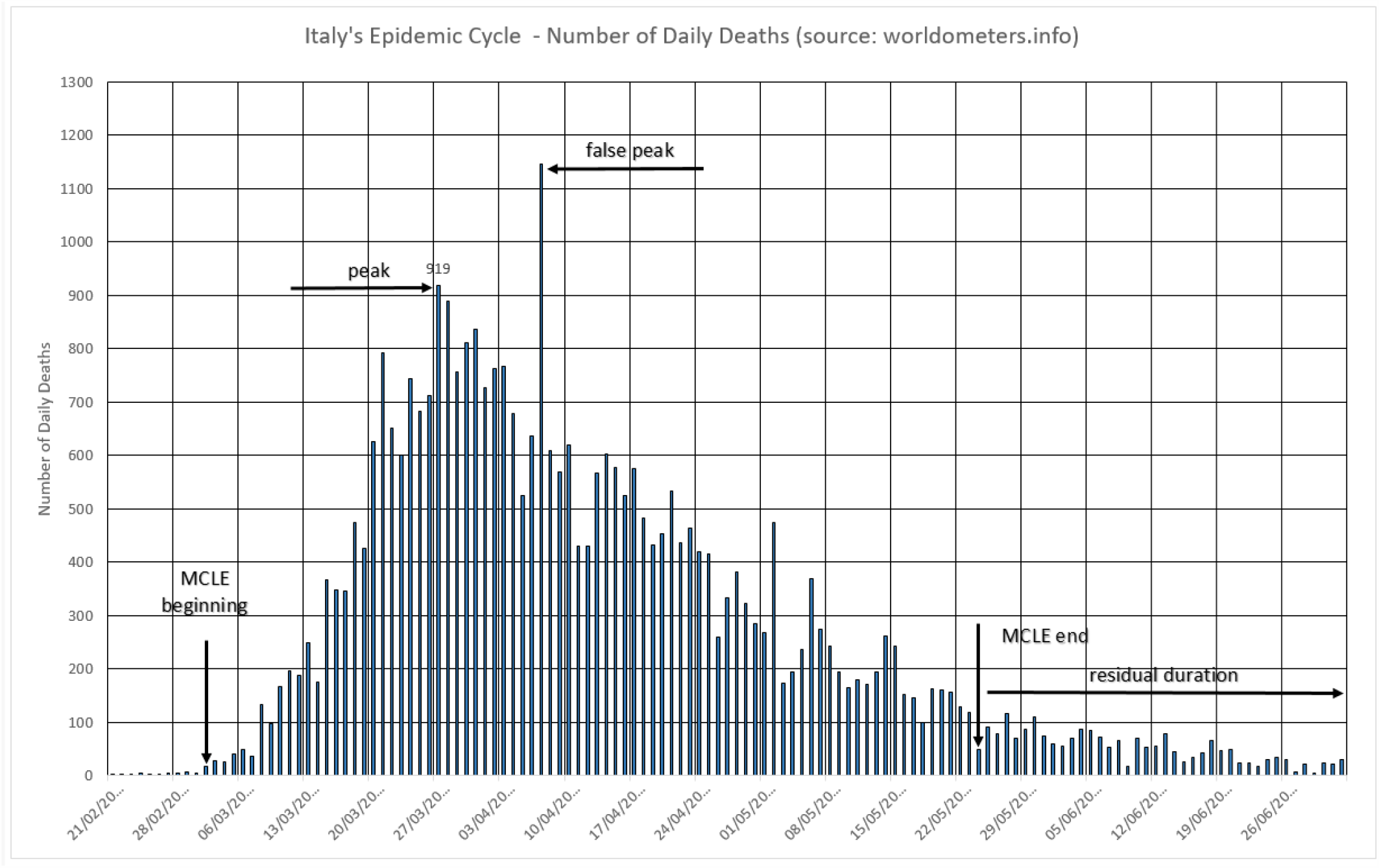
The Italian cycle.

### 4.4) New York

American city hardly hit by the epidemic, presented typical cycle found by this study. Figure 6 shows its epidemic cycle [13].

**Figure 6.**
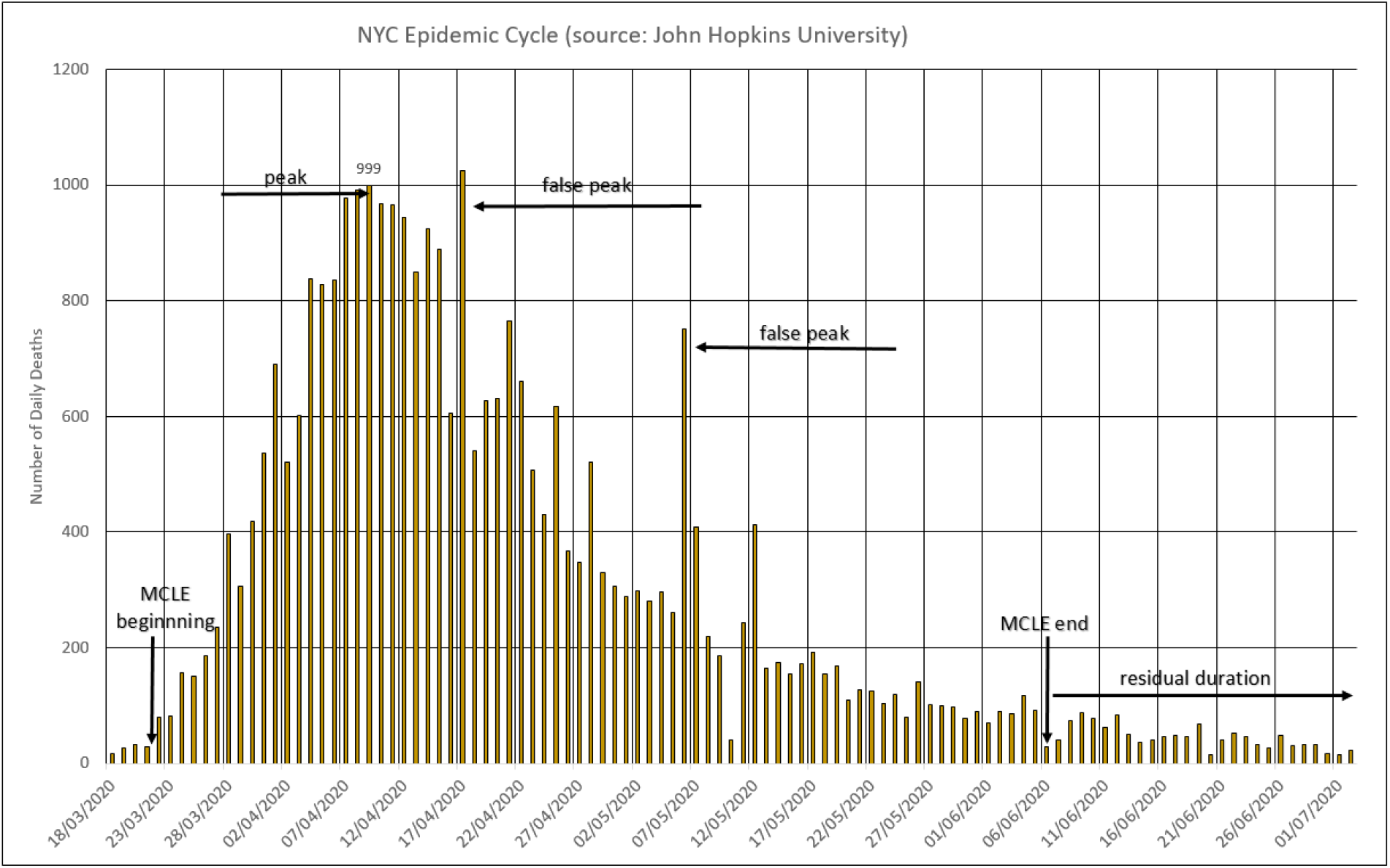
New York’s MLCE

### 4.5) Germany

Described from the beginning of the pandemic as a country that managed the crisis in an exemplary way, testing significant portions of its population and controlling and releasing the public movement based on well-known numbers and percentages of cases. Figure 7 shows the evolution of deaths in Germany [14].

**Figure 7.**
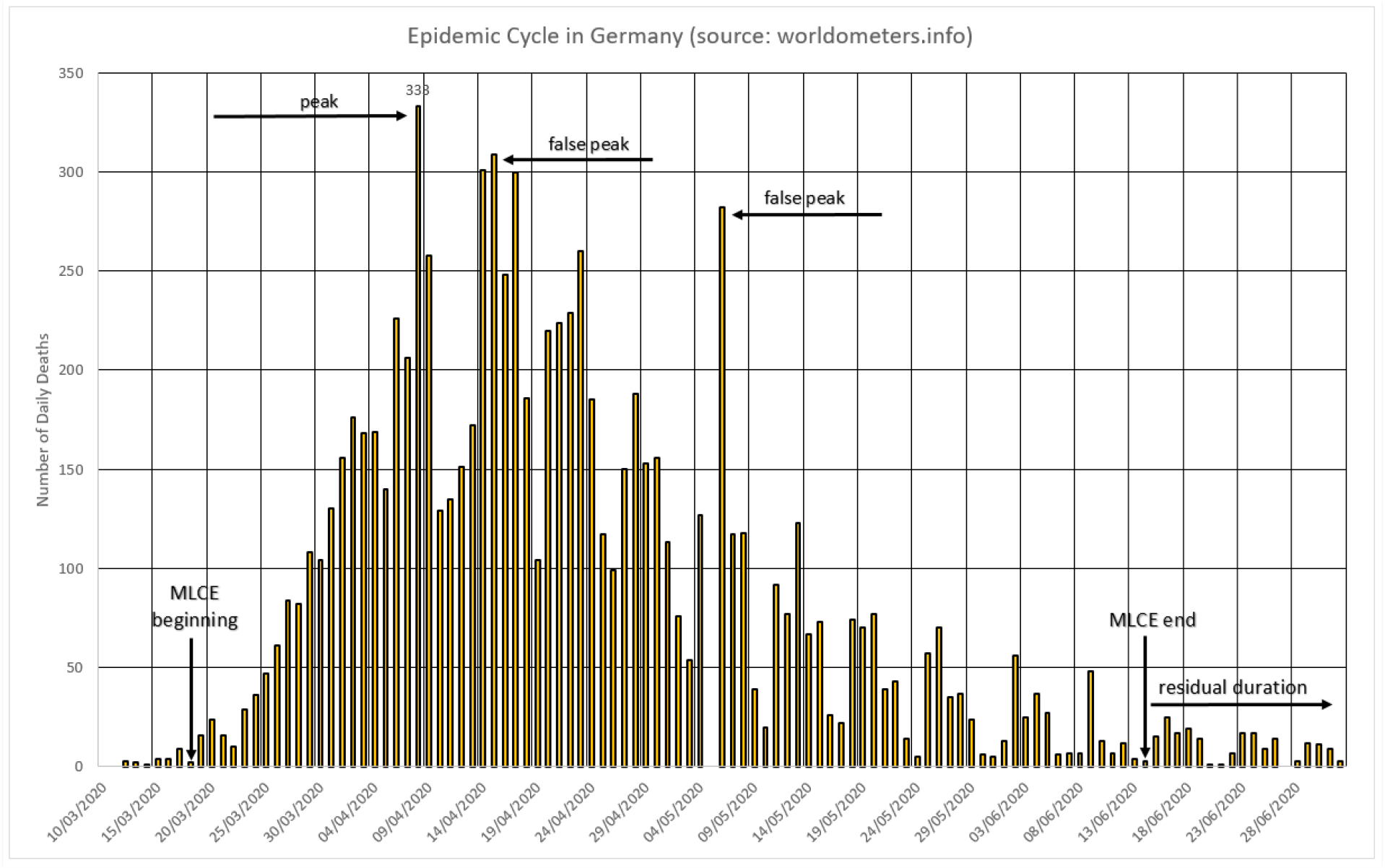
The German cycle.

### 4.6) Sweden

An European country that has not adopted the practices of radical social isolation as its neighbors, has a cycle of aspect not unlike that of all other European countries [7]. Figure 8 shows the values of deaths that have already been corrected for the dates on which they actually occurred and not the date of registration.

**Figure 8.**
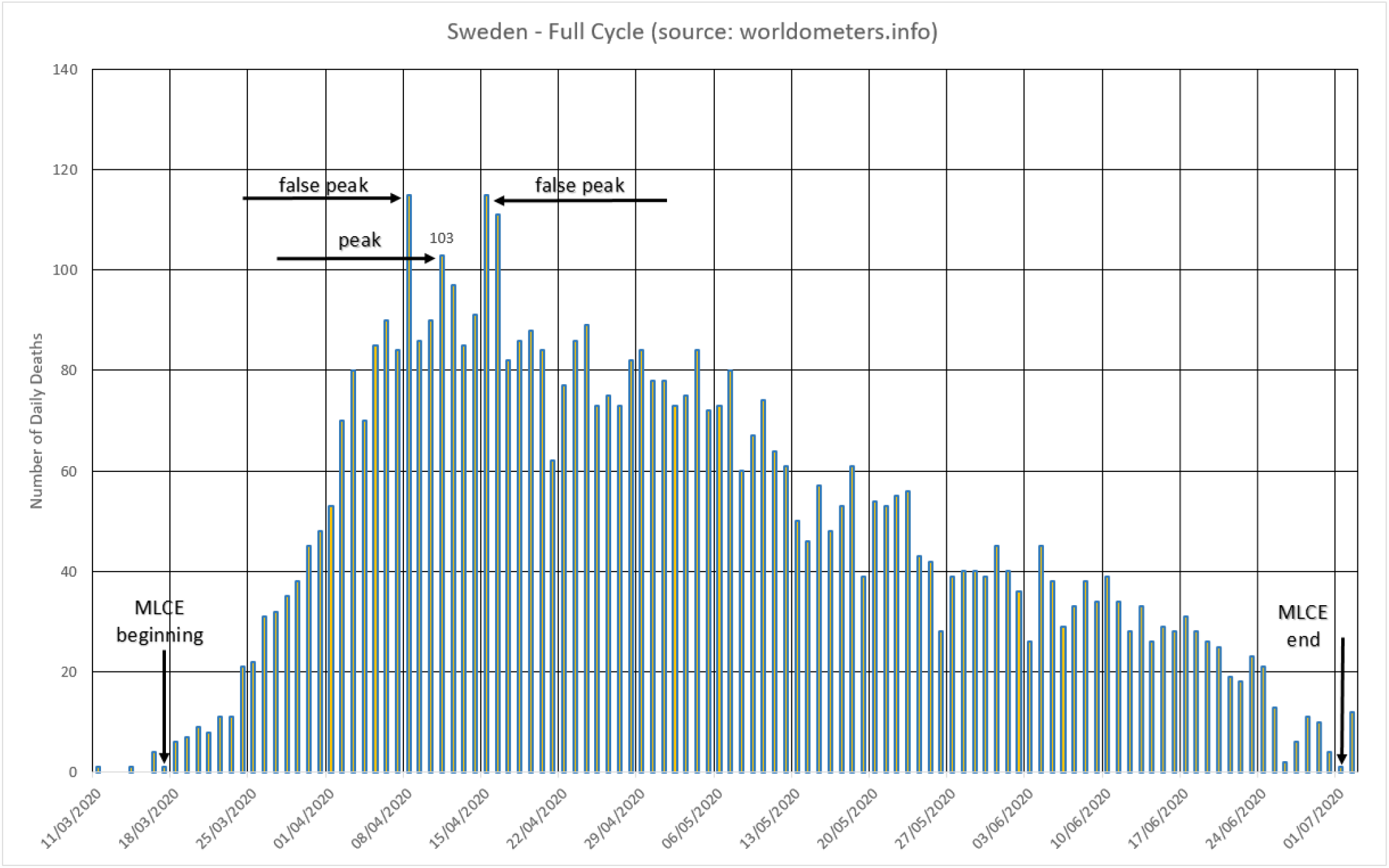
Most Lethal Cycle of the Epidemic in Sweden.

## 5) Non-dimensional Characteristics of Epidemic Cycles

In general, the epidemic cycles described here have some common geometric characteristics, the main one being a triangular aspect (Figure 9), where a smaller side is formed, which corresponds to an average daily increase in the number of deaths until a peak is reached. This peak can be easily identifiable or requires extrapolation of a line, because the values oscillate naturally and some spurious points (false peaks) may appear. After the peak, a period is formed where the number of deaths occurring daily tends to decrease on average. This period, for the observed cases, is longer than the previous one.

**Figure 9.**
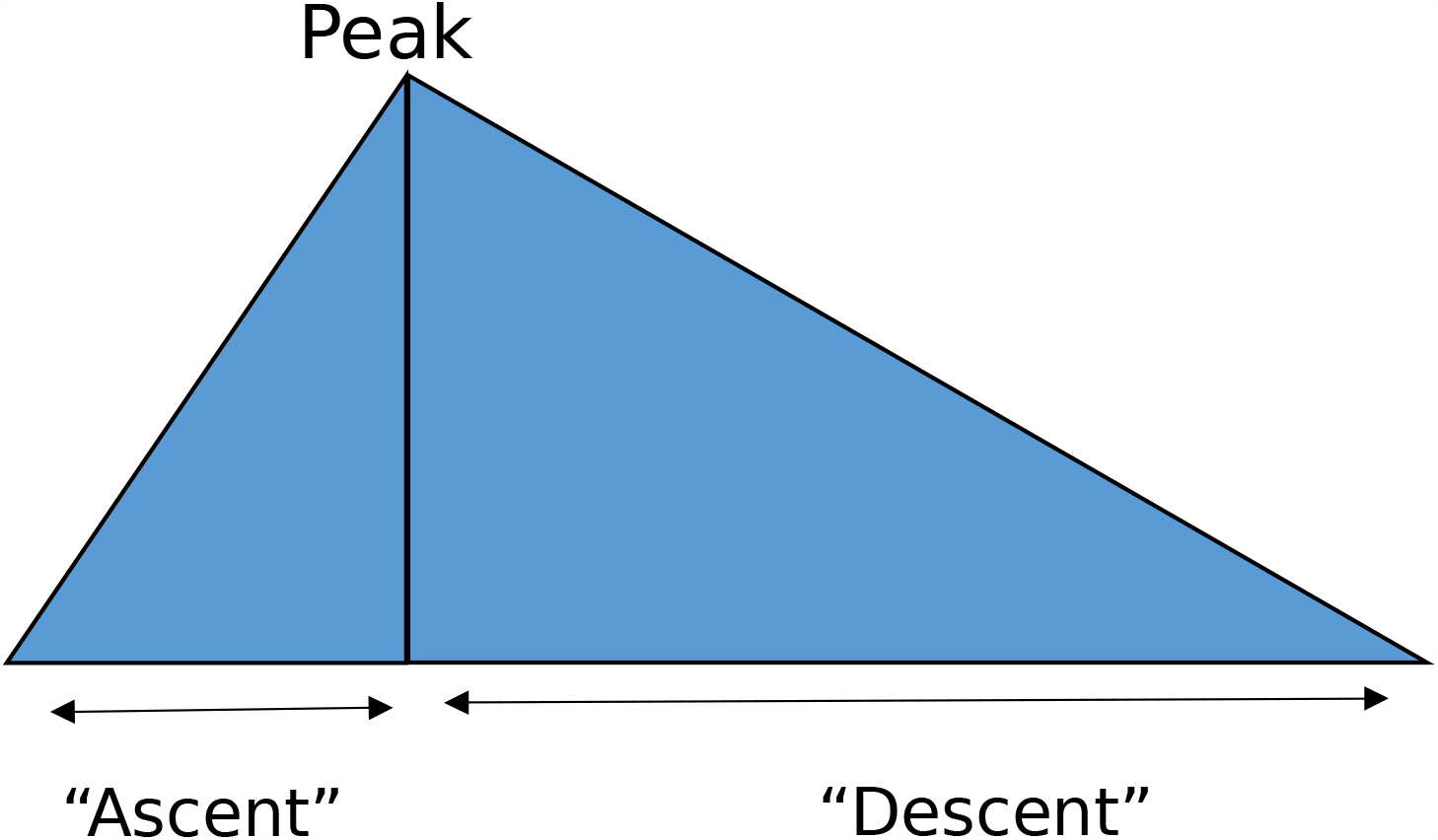
COVID-19 lethal cycles generic shape.

Hence, Table 2 gathers values of the so-called triangular cycles presented in item 4.

**Table 2.** Proportions between the time of ascent until the peak of deaths and descent until the end of the most severe cycle of the disease.

| Place | Start | Peak | End | Days to the Peak | Days to the End | Proportion between ascent and descent |
| --- | --- | --- | --- | --- | --- | --- |
| Spain | 03/12 | 04/02 | 05/18 | 21 | 45 | 2,1 |
| Italy | 03/07 | 03/27 | 05/24 | 20 | 57 | 2,9 |
| Sweden | 03/17 | 04/11 | 07/01 | 25 | 81 | 3,2 |
| Germany | 03/18 | 04/08 | 06/14 | 21 | 69 | 3,3 |
| New York | 03/21 | 04/09 | 06/06 | 19 | 55 | 2,9 |
| The Netherlands | 03/16 | 04/07 | 05/31 | 22 | 53 | 2,4 |

The values listed in Table 2 indicate that the period of rise of the disease, in countries of relatively small sizes or cities, is about 21 days, in a range that goes from 19 to 25 days until reaching the so-called peak. From then until the end of this critical period, about 60 days pass, in a range from 45 to 81 days, the ratio between the two periods oscillates between 2.1 and 3.3, with an average of 2.8. Table 3 shows the values of the number of deaths in the periods described above.

**Table 3.**
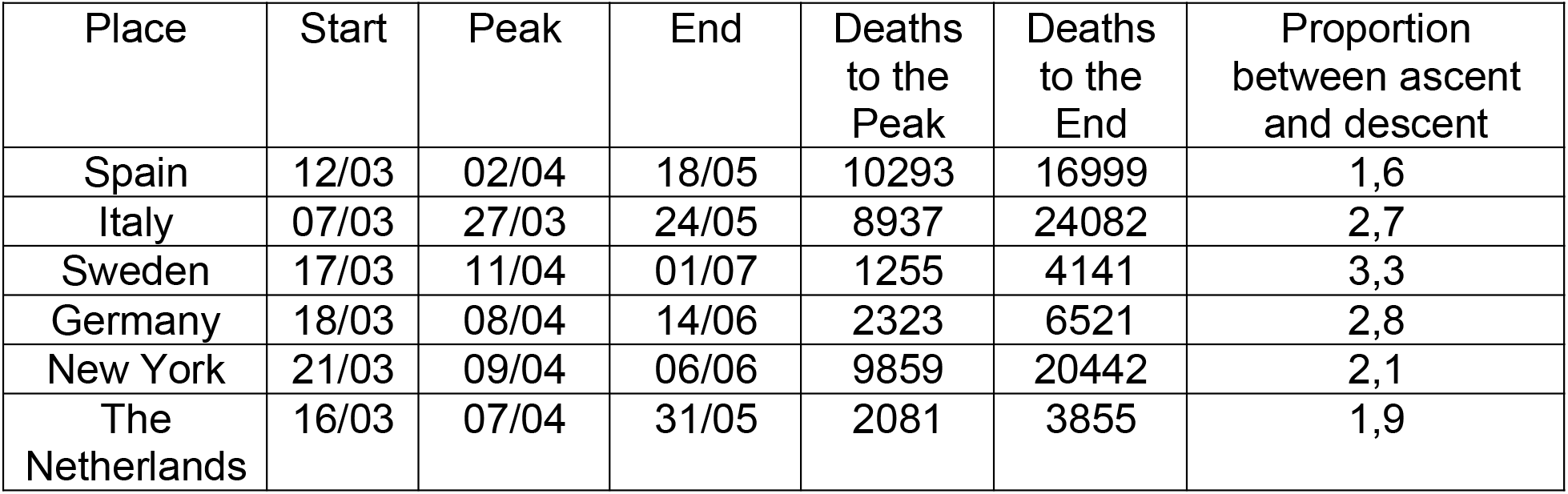
Proportions between the number of deaths associated with the cycle of rising to the peak and of descending to the end of the most severe cycle of the disease.

The values listed in Table 3 indicate that the number of deaths during the period of ascent of the disease, in countries of relatively small sizes or cities, is about 5791 deaths, in a range that goes from 1255 to 10293 deaths until reaching the peak. From then until the end of this critical period, about 12673 deaths occur, in a range from 4141 to 24082 deaths. The ratio of death figures ranges from 1.6 to 3.3, with an average of 2.4.

## 6) Similarity among Cycles

This study assumes that once the scale effects are removed, what remains is a spectrum of proportions of the epidemic cycle. Then, when submitting the data to the Moving Average Method with Initial value, there is a minimization of the effect of seasonality in the records of deaths, caused by weekends, holidays and other local peculiarities.

After dividing all the values previously transformed by the peak of the series (peak now determined by MAMI), the values start to be dimensionless and fall between 0 and 1. In this way, all epidemic cycles can be compared with each other, since what remains are the proportions between the ascent, the peak and the descent of the cycle. The time period does not change.

The hypothesis then arises that different locations may, under different behavioral rules, present the same behavior. This is what is seen next, possibly indicating that epidemic cycles are indifferent to current public health policies [15,16].

## 7) Predictability by Similarity

Some epidemic cycles observed were subjected to the numerical methods described before, and their data were subjected to a first transformation, which is the application of MAMI, Moving Average Method with Initial value, where the average of the values observed over 7 days is attributed to the first of them and so on. This allows the seasonality of death records to be mitigated. The second transformation is normalization, where all the mortality data for a given country are divided by its peak number, this will cause most of its values to be between 0 and 1 (less than the false peaks). These two consecutive transformations allow a comparison of behaviors between cycles, as well as the proof that several epidemic cycles, within the pandemic, have similarities.

With these trivial tools it is possible to estimate the duration and general behavior of a local episode, even though this, in absolute terms, does not present the same number of deaths or days of duration as its similar. What remains approximately constant are the proportions of each cycle. These are the ones that have similarities between them. An example of the application of this technique, in a very refined way, with great success is the performance prediction of professional athletes and teams made by the study group led by the mathematician Nate Silver and known as CARM-elo [17]. Below are some examples of comparisons of similar epidemic cycles.

### 7.1) Netherlands and United Kingdom

The Netherlands and the UK show a very reasonable proportion of their epidemic cycles from the beginning to the peak, during the peak of the disease and even in the initial 2/3 of the fall. The Netherlands shows a faster drop in the final part of this analyzed epidemic cycle, as can be seen in Figure 10.

**Figure 10.**
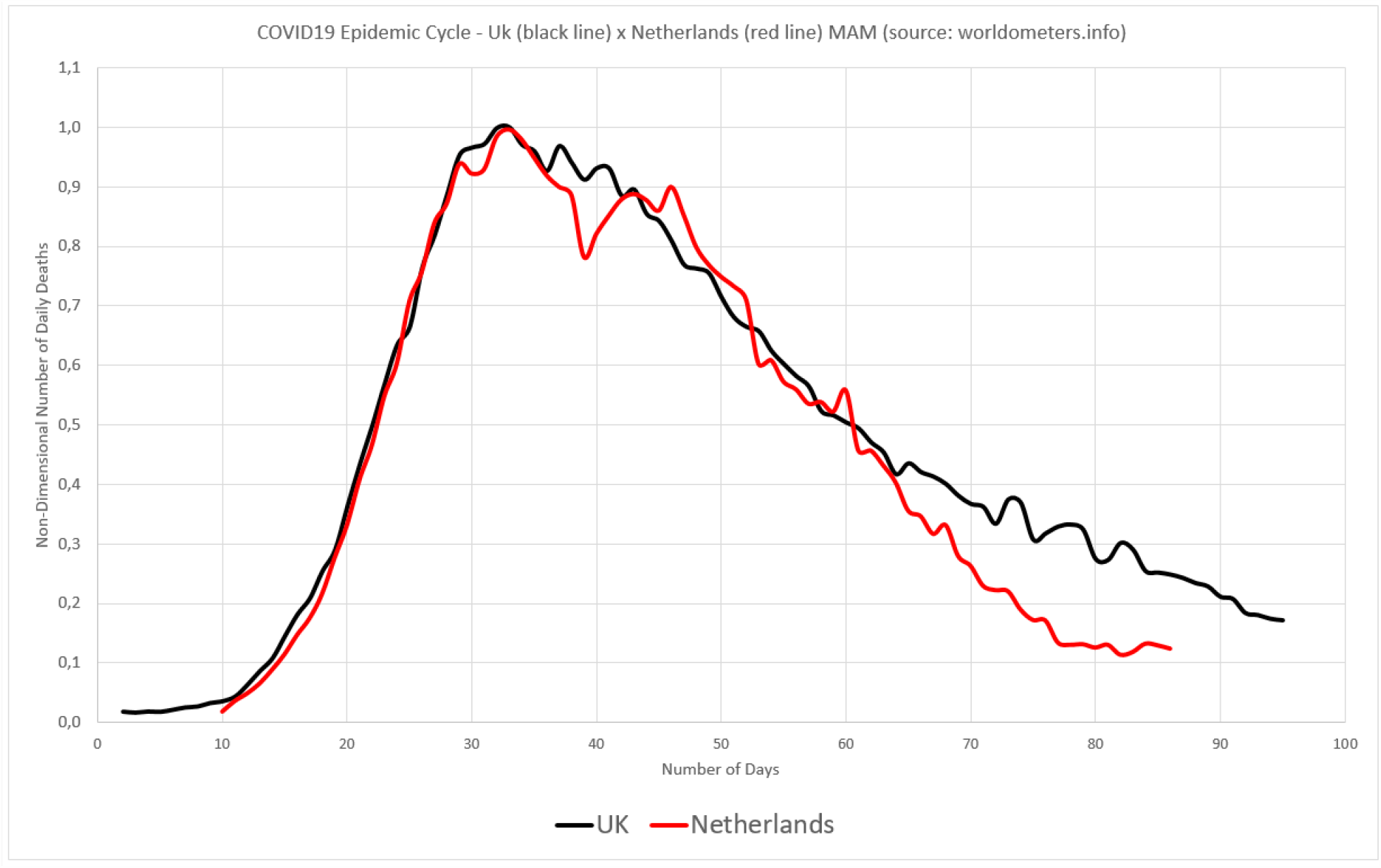
UK and Netherlands epidemic cycles compared, after their data being submitted to MAMI and non-dimension method.

### 7.2) New York and Spain

Taking into account only the proportions and not the absolute numbers, as proposed here, the great similarity between the epidemic cycles of New York City and Spain is clear. We can then observe the existence of countries that present cycles with proportions similar to that of cities, according to Figure 11.

**Figure 11.**
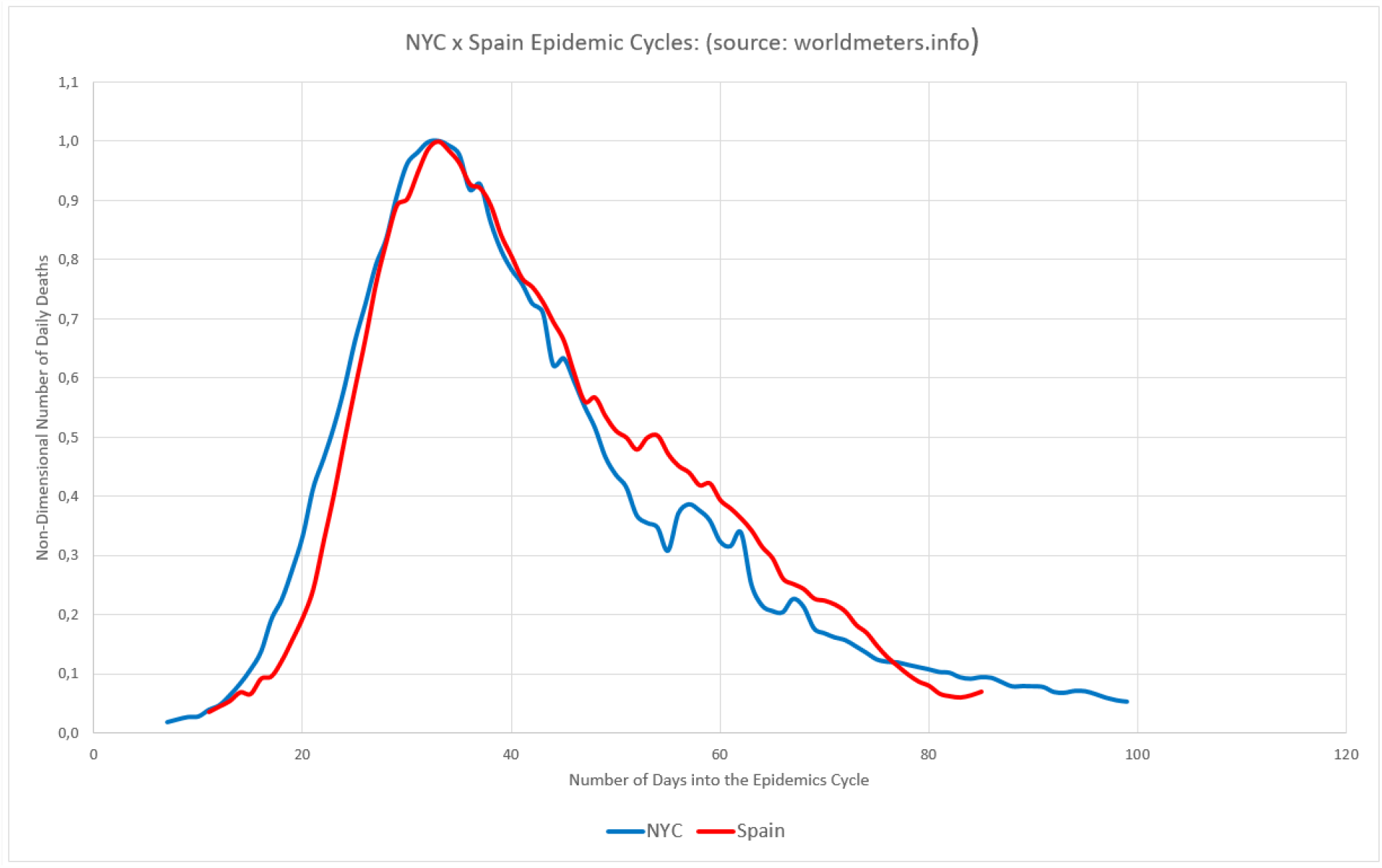
NYC and Spain epidemic cycles compared, after their data being submitted to MAMI and non-dimension method.

### 7.3) New Jersey and Belgium

Owners of very peculiar epidemic cycles, at least in the records presented, Belgium and the American state of New Jersey are quite similar when compared to each other. Figure 12.

**Figure 12.**
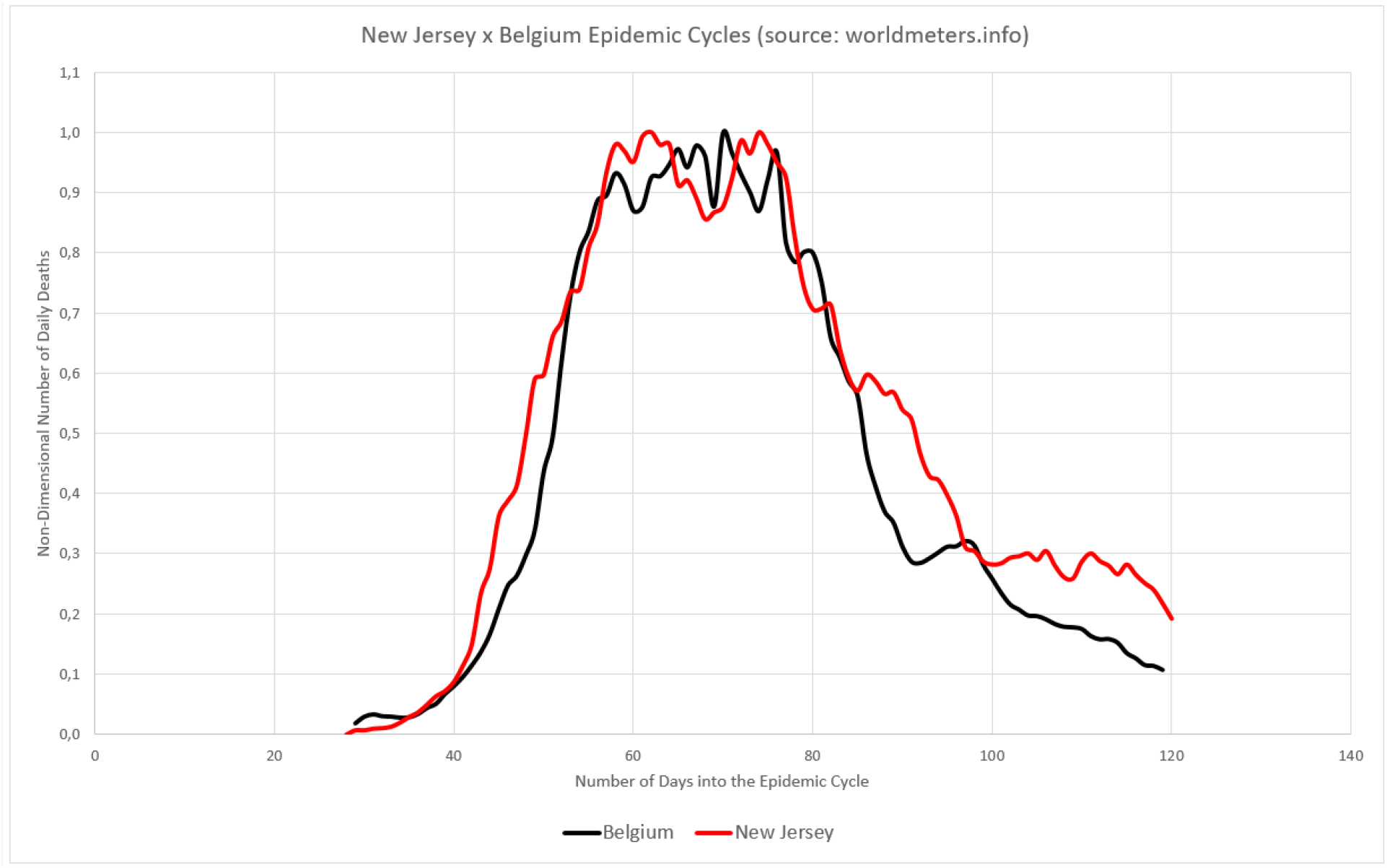
Belgium and New Jersey epidemic cycles compared, after their data being submitted to MAMI and non-dimension method.

**Figure 13.**
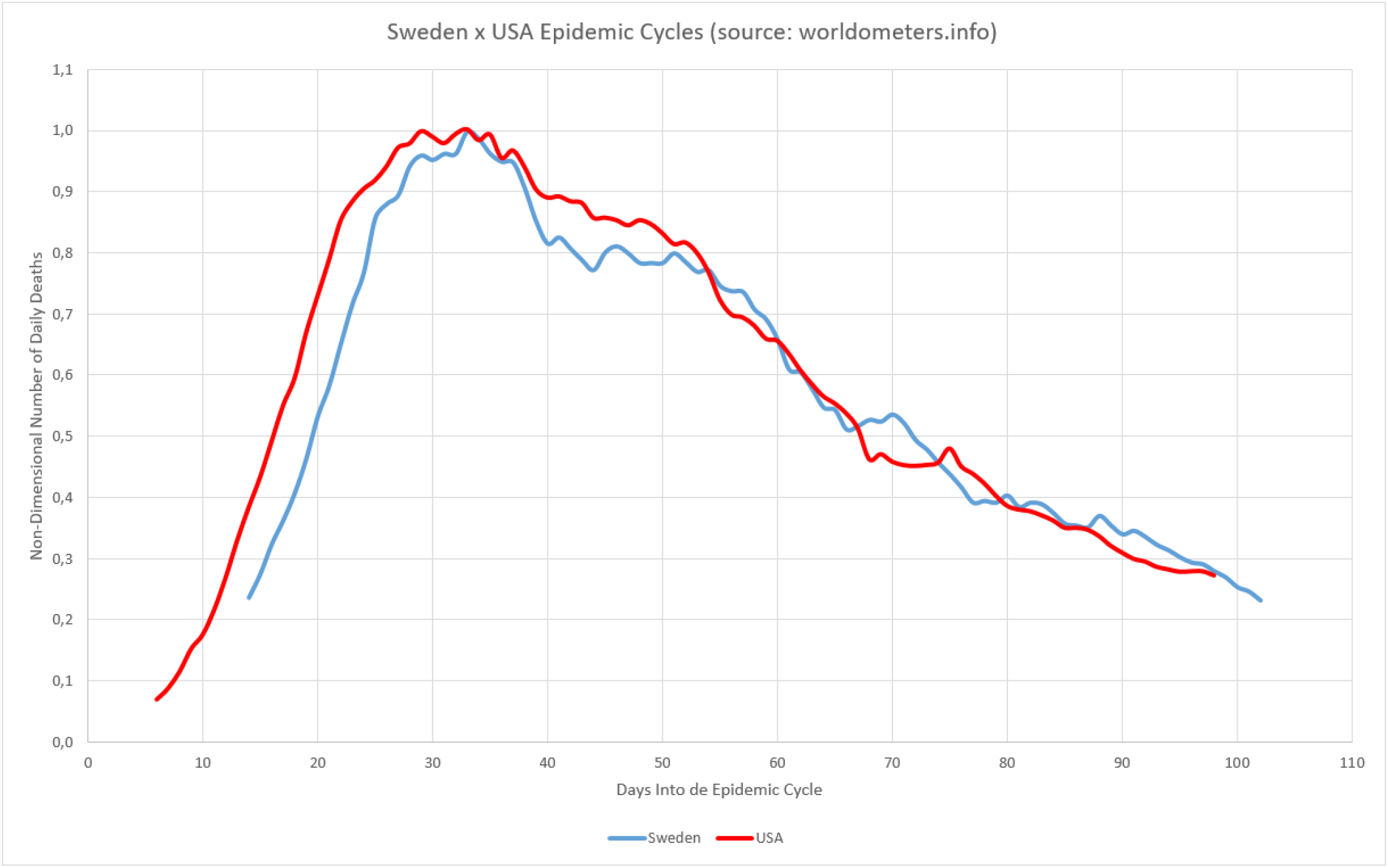
Sweden and USA epidemic cycles compared, after their data being submitted to MAMI and non-dimension method.

### 7.4) USA and Sweden

Completely different countries in regard to their territorial extensions, human complexity, geographical position etc. However, the proportions in the evolution of their critical epidemic cycles are very similar.

### 7.5) Sweden x Germany

It is important to note that, to use prediction by similarity, it is necessary to stress that the cycle used as the basis for the prediction has ended - in the terms proposed here, since the fact that cycles show similarity in the ascent does not guarantee that they will show similar descents. To illustrate this situation, a comparison follows between Sweden and Germany, where not only in the ascent, but throughout the critical period, the two countries had very similar behaviors. However, the descent has shown varying degrees for the two countries.

Figure 14 presents a fully normalized comparison of the Swedish cycle with the German cycle. In Zone A, the Swedish cycle has a very similar behavior and at times less aggressive than the German one, while it starts to detach from the second in Zone B, it moves further away in C, and finally converges in D at the end of the cycles. This is a point to be studied: the pattern indicates that at the peak of the Swedish cases (with 51% of deaths in Zone A), the behavior was the same as that of the German cycle, but the Swedish decent part of the cycle was slower.

**Figure 14.**
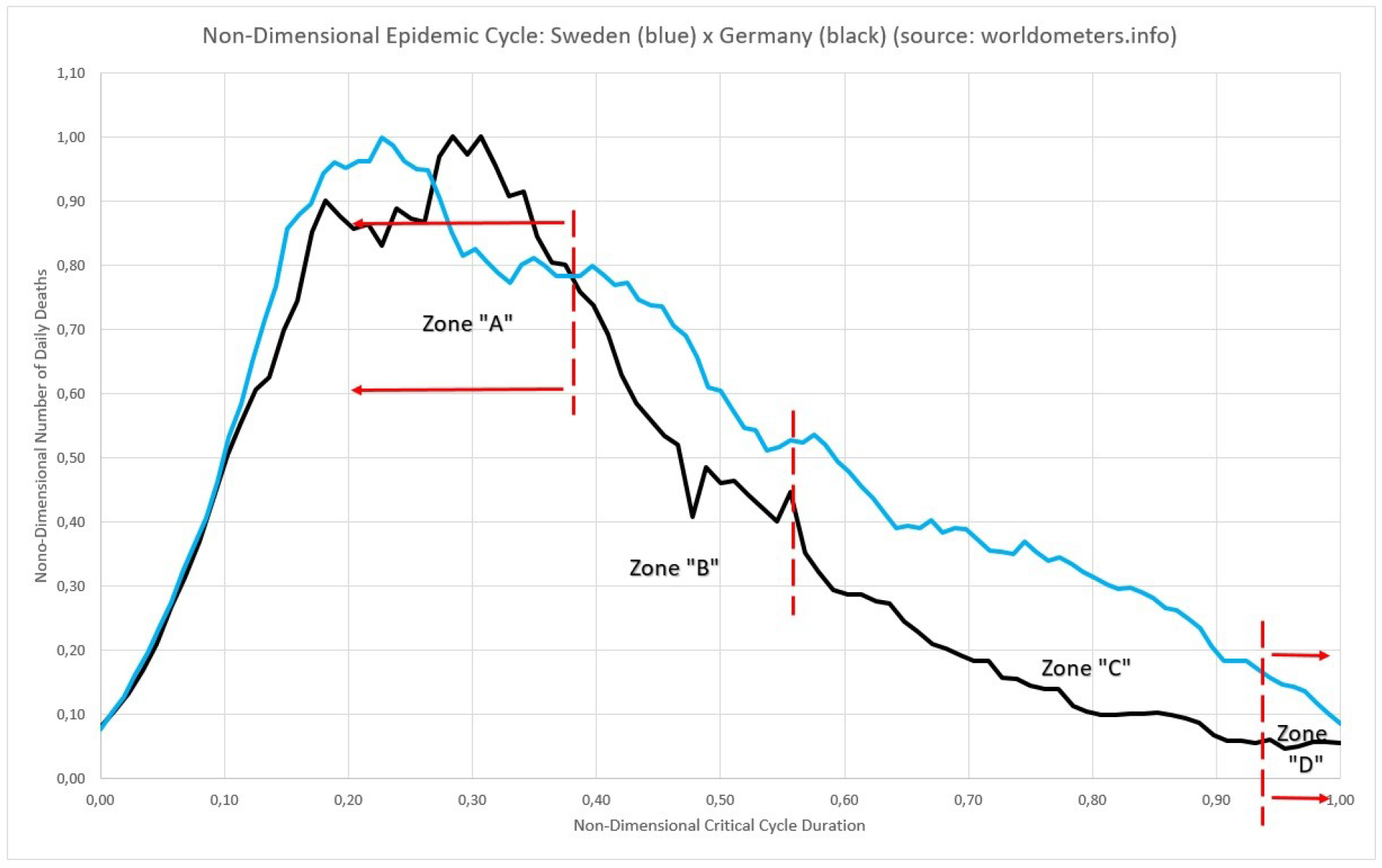
Fully Normalized Sweden X Germany epidemic cycles compared.

## 8) Brazil, State of Rio de Janeiro and the City of Rio de Janeiro

Brazil, if seen as a whole, has the same slow cycle of rise as India and Mexico; and a long cycle like the USA. They are all countries with large territorial extensions and population concentrations. Russia, Australia and Canada are large countries, but with low population density, therefore with another type of epidemic cycle. The authors understand that countries like Brazil should be seen and dealt in terms of counties or regions, with regard to the study of epidemic cycles, since regions that started their cycles at the beginning of the pandemic, are already at the end or close to it, and those who have only had residual cases so far can start the acute cycle at any time. Seen as a whole they seem to be a long cycle and different from all those presented here in this work, but if seen individually, they are quite similar to the “triangular” cycles presented in topic 4. Countries with a large population and continental dimensions, may have their epidemic cycle curve built from the convolution of the curves of the cities with the largest population.

The data sources used in this study for Brazil are the Ministry of Health’s websites [18] and National Council for Justice’s Transparency Portal [19]. The Ministry of Health (MoH) publishes data on deaths on the registration dates, in the same way as several countries in the world presented here, implying the appearance of the inevitable “saw” aspect in the graphics. The second site presents the number of deaths on the dates when they actually occurred, but inevitably suffer from a delay in updating values, due to the time required for family members to register death and the information to become part of the system. The numerical tools presented minimize the effects described and allow a clearer view of the epidemic in Brazil.

### 8.1) Ministry of Health

The Ministry of Health publishes in a daily basis the sum of the number of deaths received and verified, from the state health departments and the Federal District. These are the numbers recorded that day. The death may have happened many days before, but it will be counted on the date of registration. The effect of this is sharp drops on weekends and extreme peaks in the middle of the weeks. But their numbers reflect the average (MAMI) of the true values of deaths in the country. If there are adulterations of values, this occurs in the origins of the values and not in these totals, therefore this work will consider this source to be valid. Confirming the validity of this source, there is the following reference, whose values are very similar to those of the MoH and tend to converge over time, implying not only the veracity of the figures presented, as well as the absence of significant underreporting in the MoH numbers.

### 8.2) Transparency Portal (Portal da Transparência)

Presented as an initiative by the Brazilian Civil Registry Offices, this website compiles death registration data across the country, with special attention to COVID-19 data. The data is updated about three times a day. It is delayed in relation to the real value, due to the obvious need of families to register deaths on dates after their events. Its values for cities like Rio de Janeiro vary markedly only in the most recent days (up to 4 days on average), however considering Brazil as a whole and its more distant regions, this variation occurs for a longer time. Thus, there are two variations to be observed in this database, the first being how long it takes a given day on the calendar to reach a given value that will be considered to have “converged”, that is: from a given date it will only will change residually, not affecting any study done, the same can be seen in the international database referenced here [7]. The second variation stems from the fact that the calendar continues normally and the days advance, with new deaths being launched and the most recent days are, by definition, underreported. This work evaluated how long it takes for a given value to reach stability, which was defined as being when it stopped varying more than 1% within a week. Appendix A presents a comparative study between the two portals.

#### 8.2.1) Time to Value Convergence - City of Rio de Janeiro

In order to assess the time it takes a certain death curve to reach stability, with stability defined here as that level that varies less than 10%, the values of the city of Rio de Janeiro were monitored. Evaluating the graph in Figure 15, the first observation is that only the last 4 days vary by more than 10% over the course of a week. In other words: after 2 weeks there is no more significant change in the values presented, except for the last 4 days. From the first day (blue, 23 of June) to the second day observed (29 of June), 6 days pass and although on average the values are less than 10% (6.0 calculated), there are isolated cases that varied more than what is this. However, between 06/29 and 07/07, 6 days pass again and now the average variation is 0.3%, if the last 4 days are excluded. Thus, it can be said that for the city of Rio de Janeiro, currently, it is enough to eliminate the last 4 days to make an accurate projection of the evolution of the epidemic cycle in the city. In the same way, you can study and evaluate any and all cases covered by the Transparency website.

**Figure 15.**
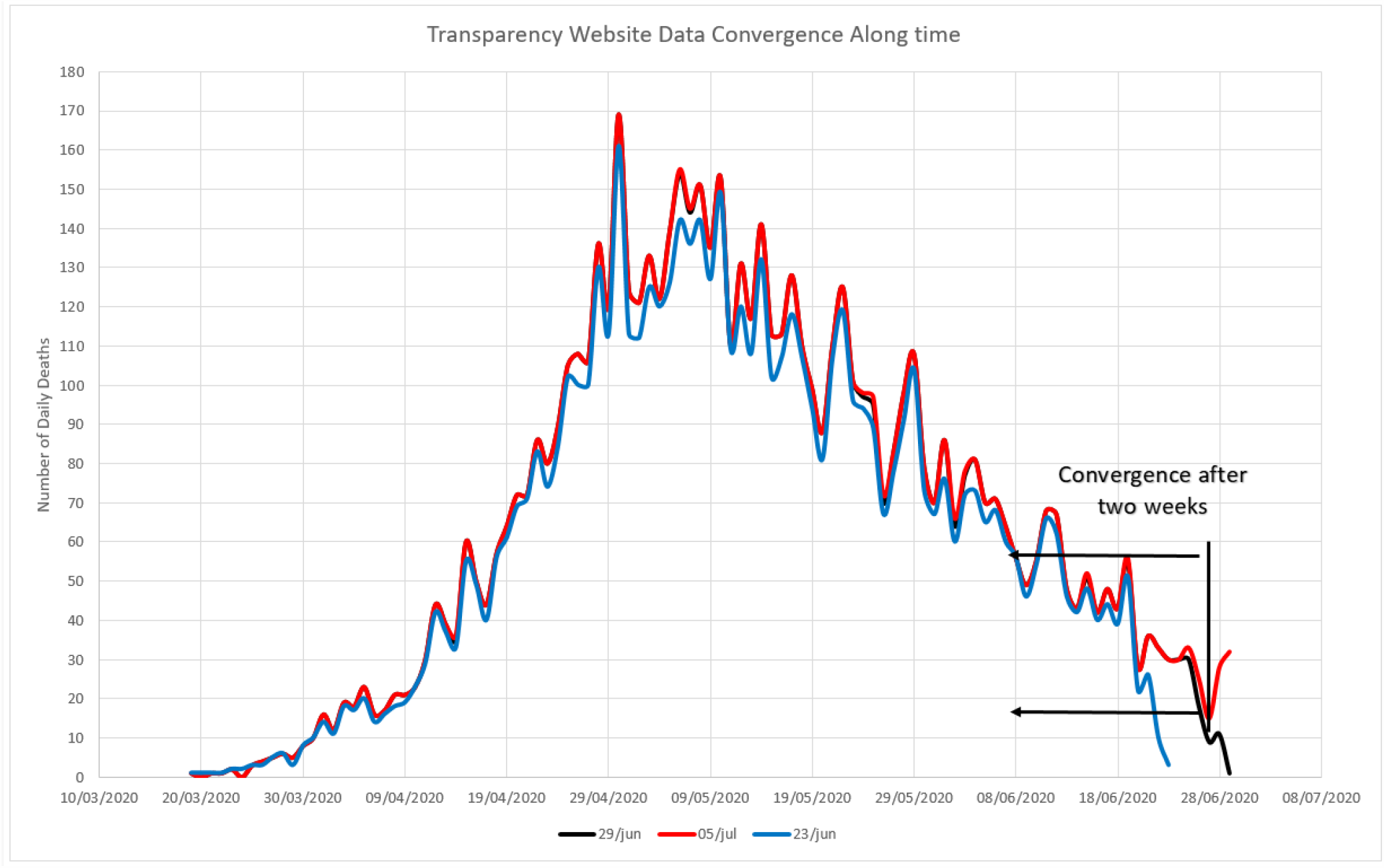
Daily death figures data for City of Rio de Janeiro, in three days. There are no noticeable difference between the second and the third weeks, only the last four days.

#### 8.2.1) Time to Value Convergence - Brazil

This work considered that variations in the number of deaths in a given day less than 5% are negligible and that up to 10% can be accepted without elimination. This implies that the data to be deleted will be that of the last 10 days. This can be seen in Figure 16. Comparing the data from the 5th of July (black line) to the 29th of June (red line) it is noted that before that, from the 17th day backwards, the values are below 3% and in average 1%, all with no practical effect on any projection you want to make (Zone A). Even when observing the values of June 24 (blue curve), although the variation occurs above 10% in a period over 10 days (Zone B), the shape and proportions of the curve are identical to the two curves on dates later, this indicates that the distribution of later notifications does not change the shape of the curves and that if they are subjected to the numerical methods mentioned above, they can in fact serve predictions with acceptable accuracy.

**Figure 16.**
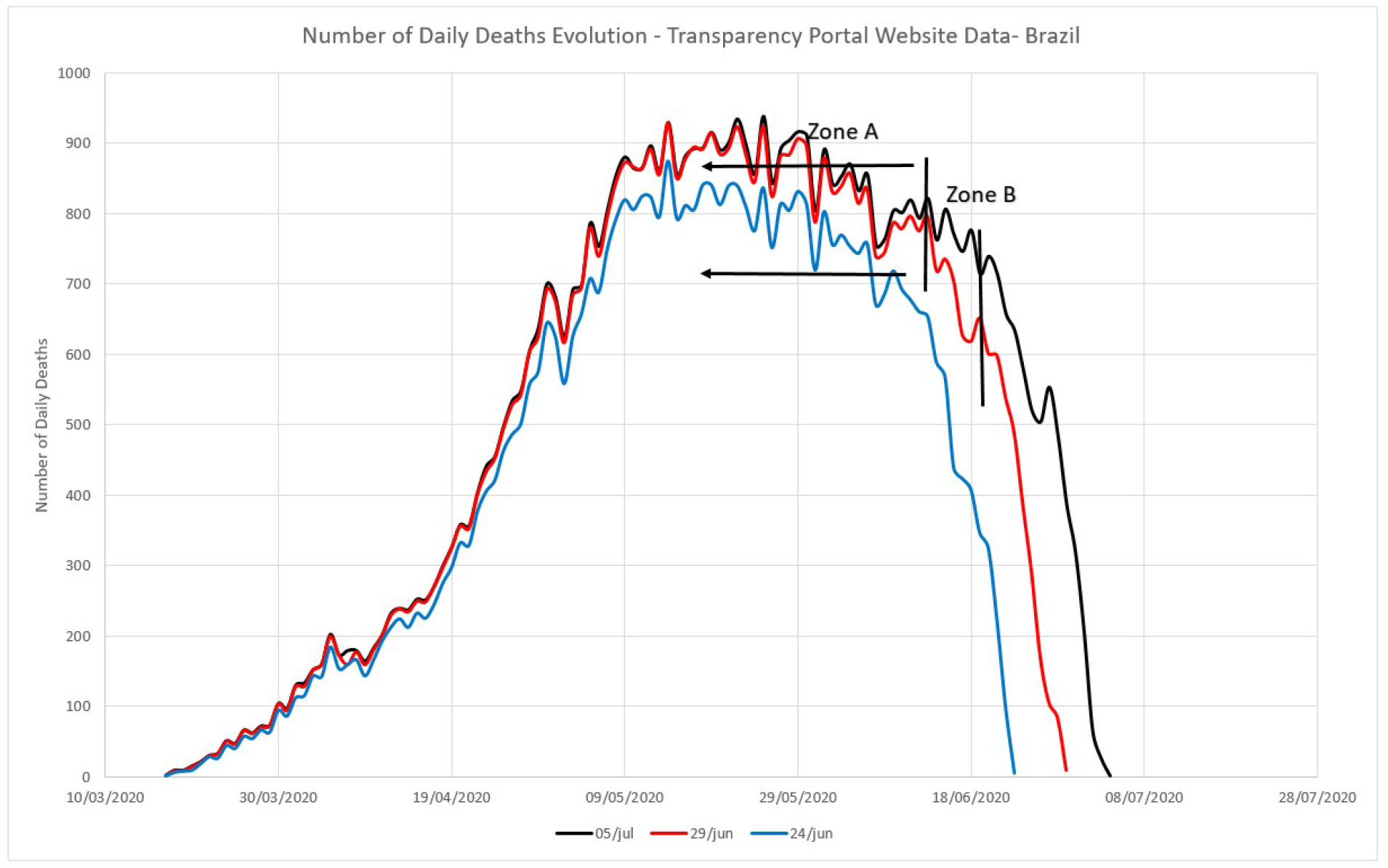
Daily death figures data for Brazil, in three days. There are no noticeable difference between the second and the third weeks, only the last ten days.

**Figure 17.**
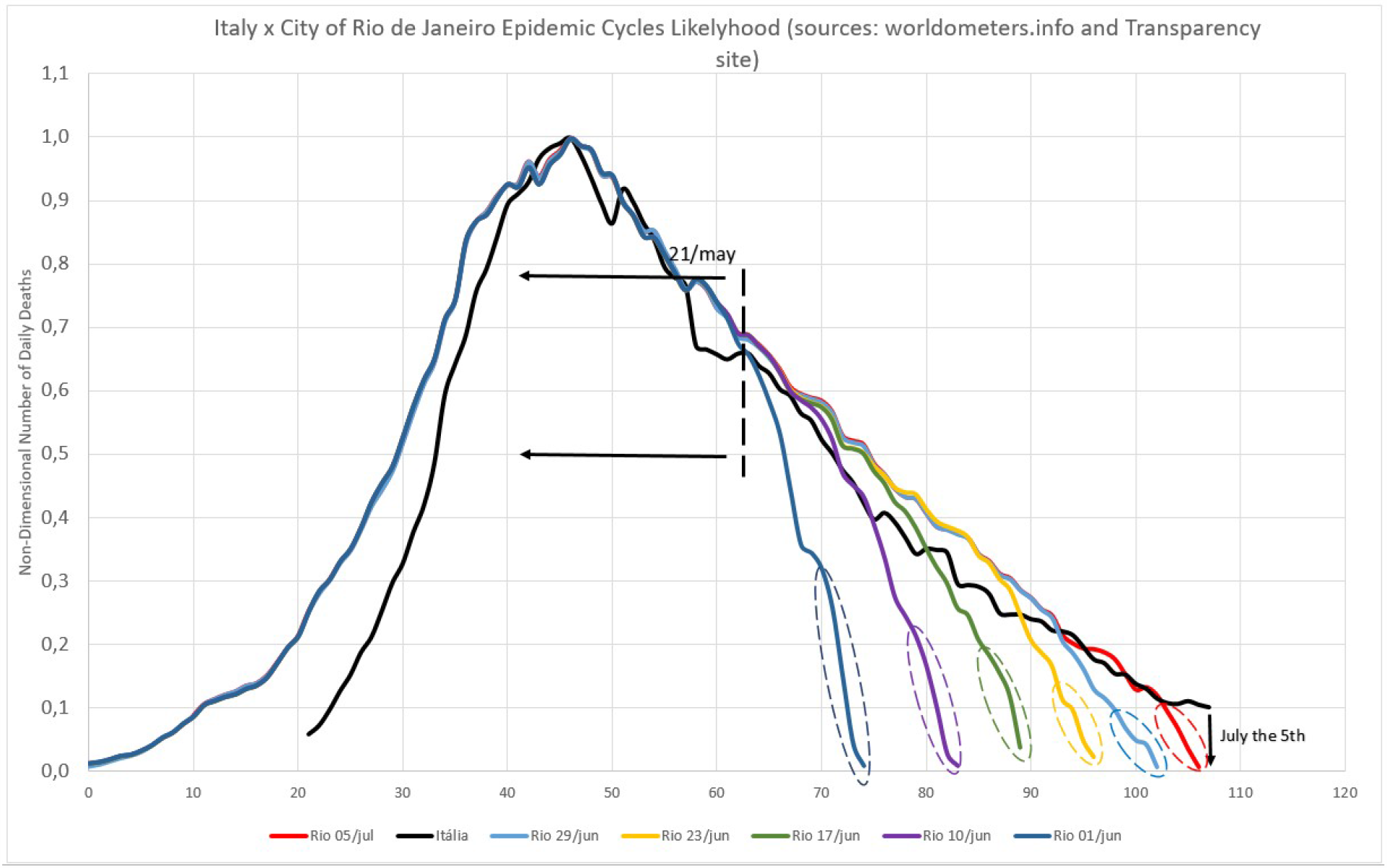
Several evaluations for the end of the cycle comparing the city of Rio de Janeiro and Italy.

### 8.3) Total Deaths

#### 8.3.1) Brazil

The inevitable delay between the occurrence of physical death and the registry at the registry office has an effect on the count of total deaths in Brazil and in each location. At the end of the epidemic the two values (MoH and Transparency Portal will converge, but during the outbreak, there will always be a delay between the two). The following example was carried out on the afternoon of July 5, 2020. The total number of deaths indicated by the Transparency Portal website was 58155, while the MoH website had the number 64265 deaths. A difference of 6110 deaths (9.5% of the total) that occurred, but have not yet been registered.

#### 8.3.2) City of Rio de Janeiro

In the case of the City of Rio de Janeiro, for data collected on the morning of July 6, 2020, on the two websites mentioned, it reveals that the Ministry of Health points to a total of 6898 deaths in the city so far and the Transparency Portal (TP) indicates 6855, a difference of 43 deaths, that is, about 0.6%, indicating an excellent coherence between the two total values, and the distribution, after the application of MAMI, indicates the existence of the convertibility of the MoH values to the TP, although not as complete as in the case of Sweden, previously described.

### 9) Cycle Predictions in Brazil

From all the numerical tools and the databases presented, as well as the limitations and possibilities in their use, the authors are now making projections for the temporal behavior of epidemic cycles and the number of deaths associated with them. Several forecasts presented here have already been released on social networks on different dates in the form of postings and so-called “lives” [20], being available on the platforms for consultations and confirmations.

Predictions are made following a very simple script:

a. The daily death data for a given location are noted in a spreadsheet and the MAMI calculated.
b. The values are normalized, as already explained, the MAMI is applied, because although the registry office records deaths on the correct dates, so that the two data series undergo the same numerical transformations.
c. A continuous curve is generated on a graph with the x-axis as the number of consecutive days of the epidemic cycle and the y-axis, the dimensionless range from 0 to 1, generally (some points, the false peaks, can go beyond this).
d. Among countries or localities, we seek those that have already ended their critical epidemic cycle (which was called in this work MLCE) and that is visually similar to the curve obtained in (c), although obviously on a different scale.
e. MAMI is applied.
f. Data is normalized.
g. Repeat (c).
h. As the country or locality of reference has already finished its cycle, it will be positioned previously on the graph, in relation to the place where it is desired to estimate the probable end date of the critical cycle. You should then numerically superimpose the peak of the case you want to study, for example, with the reference.
i. Once the superposition is made, always moving the reference case, an extrapolation can be made using the reference case as a guide to the value to be determined. As the scale of the case studied has not been changed, it is enough to consult what day it would be in the future to know the probable date.
j. If there is no similar case, you can eliminate the last days, as discussed above, and extrapolate directly from the values obtained in the Transparency Portal database.

#### 9.1) Rio de Janeiro City Cycle Prediction

The forecast of the cycle of the city of Rio de Janeiro will be carried out by the two methods proposed here, 1) the superposition of a cycle that has already ended, in this case Italy, with that of the city and 2) the extrapolation from the data series, with the elimination of the last values.

#### 9.1.1) City of Rio de Janeiro and Italy

Since Italy was the case chosen to serve as a forecasting model, the predicted values were updated based on the daily values obtained on the TP website, that is: values presented here were collected on the mentioned dates. Although the cycle of climb in Italy was not similar to that of the city of Rio de Janeiro, the descent cycle was considered to be good enough in the initial moments and this model was adopted, which proved to be correct. It was considered in item 4 that the Italian critical cycle ended when the level of daily deaths decreased until reaching 10.6% of the peak already submitted to MAMI, from then on the residual cycle started. For the city of Rio de Janeiro, after the MAMI, the peak value becomes 143, so 10.6% of this would be 15 deaths. So when the city of Rio de Janeiro reaches approximately 15 daily deaths, the critical cycle can be considered closed. Note that on the forecast day, July 5, the curves met.

In the first assessment, made on June 1, 2020 (dark blue curve), the concordance between the proportions graph of Italy (black curve) and the city of Rio de Janeiro coincides until May 21, after which there is a great divergence, but it is possible to note that: a) the cycle is still far from the end and b) the proposed cycle describes the critical episode in the deceleration well, for about 1/3 of the period. If the appraiser accepts this period as being sufficient to characterize a valid similarity, it is sufficient to consult the final value of the Italian episode (10.6%) and estimate the date when the Rio de Janeiro’s cycle will reach that value of its peak, as described above.

On June 10 (purple curve) as well as on all dates used in this graph for the city of Rio de Janeiro, the last 4 days were marked, which are contained within the ellipses. Observing the data of these 4 final dates of each series guarantees that they will grow and, therefore, become even more similar to the Italian model, as is evident when analyzing the later dates, but at the time of collection they were still at their minimum. The agreement on this curve runs until June 2, then diverges, but more than half of the descent cycle agrees with the Italian, so the forecast becomes quite assertive.

For the data collected on June 17 (green curve), the curve agreement runs until June 7, then diverges slightly until June 10, but the agreement with the Italian is quite obvious and the date foreseen for this cycle, 05 July, is increasingly confirmed.

For June 23 (yellow curve), the data coincides until June 16 and the whole curve agrees on more than 4/5 of its extension, confirming the initial estimate of July 5.

On 06/29 (blue curve), the data are intertwined with the Italian cycle until June 19, always confirming initial estimates.

On 07/05 (red curve) there is a strong interweaving of this data, confirming the end of the cycle by this estimate. It remains to wait for the evolution of the disease in the city and to compare the estimate with the real values.

The critical part of the fall cycle in the city of Rio de Janeiro was then estimated to have started on May 4, 2020 and ended on July 5, where it would have reached the value where the residual cycle begins. Thus, the fall cycle lasted 62 days. Joining the 42 climb, it is 104 days in total, that is, a cycle of almost 15 weeks.

Table 4 below compiles the values mentioned above.

**Table 4.** Comparative values determined by Italy and City of Rio de Janeiro epidemic cycles.

| Data Collection Day | Limit of Good Agreement With Italy | Total Days of Good Agreement | Good Agreement and Total Descent Cycle Ratio | Days to the End of the Predicted Epidemic Cycle |
| --- | --- | --- | --- | --- |
| 01/jun | 21/may/2020 | 17 | 0,27 | 34 |
| 10/jun | 02/jun/2020 | 29 | 0,47 | 25 |
| 17/jun | 07/jun/2020 | 44 | 0,71 | 18 |
| 23/jun | 16/jun/2020 | 50 | 0,81 | 12 |
| 29/jun | 19jun/2020 | 56 | 0,90 | 6 |
| 05/jul | 30/jun/2020 | 57 | 0,92 | 5 |

The values in Table 4 indicate that it was already possible to make an accurate estimate for the end of the critical epidemic cycle in the city of Rio de Janeiro on June 1 (35 days before the end), although this contained a greater degree of uncertainty, since only 29% of the descent curve agreed. However, as of June 10 (25 days in advance), with 47% agreement between the curves, the date could already be estimated. As of June 17, there is a very strong agreement between the curves, above 70% and the forecast becomes even safer. If the last 4 days of the series are discarded, as already discussed, it can be noted that the series of July 5 becomes identical to that of Italy from June 30.

#### 9.1.2) Data Extrapolation

To serve as an example, three dates and data that were collected on those dates on the TP website are selected. The data collection dates were 06/23, 06/29 and 07/05/2020. After this, a selection of the peak is made as the last point at which the series rose and from then on it started to decrease, even if in an irregular way. The points (including this peak) are selected excluding the last 4, for the reasons already discussed, submit them to MAMI and apply the linear regression process, where a line is forced to pass through these points. The equation for this line is then equaled to the null value and then the number of days of the epidemic cycle is found, with Day Zero being March 19, 2020, the date of the first death that occurred, which corresponds to a date in the spreadsheet. The values obtained are listed in Table 5, at the end of this item. Figure 18 presents the example of the case for the data collected on July 5, 2020. All other cases presented here were calculated in the same way, as already described. It can be seen from the data in Table 5 that the time (in days) between the day the data was observed and the predicted date, is reduced as the end of the cycle approaches. This is achieved when the difference between the predicted day and the day of the data used is zero.

**Table 5.** Estimation results for several different days.

| Day | Data Fitting Eqn. | R<sup>2</sup> | Days into the cycle | Estimated Day | Days to the end of the cycle |
| --- | --- | --- | --- | --- | --- |
| 23/06 | $y = -2,2263x + 289,68$ | 0,9939 | 104 | 01/07/2020 | 8 |
| 29/06 | $y = -2,3338x + 304,76$ | 0,9961 | 105 | 02/07/2020 | 3 |
| 05/07 | $y = -2,2086x + 237,70$ | 0,9951 | 107 | 05/07/2020 | 1 |
| 07/07 | $y = -2,1864x + 236,24$ | 0,9933 | 108 | 06/07/2020 | 0 |

**Figure 18.**
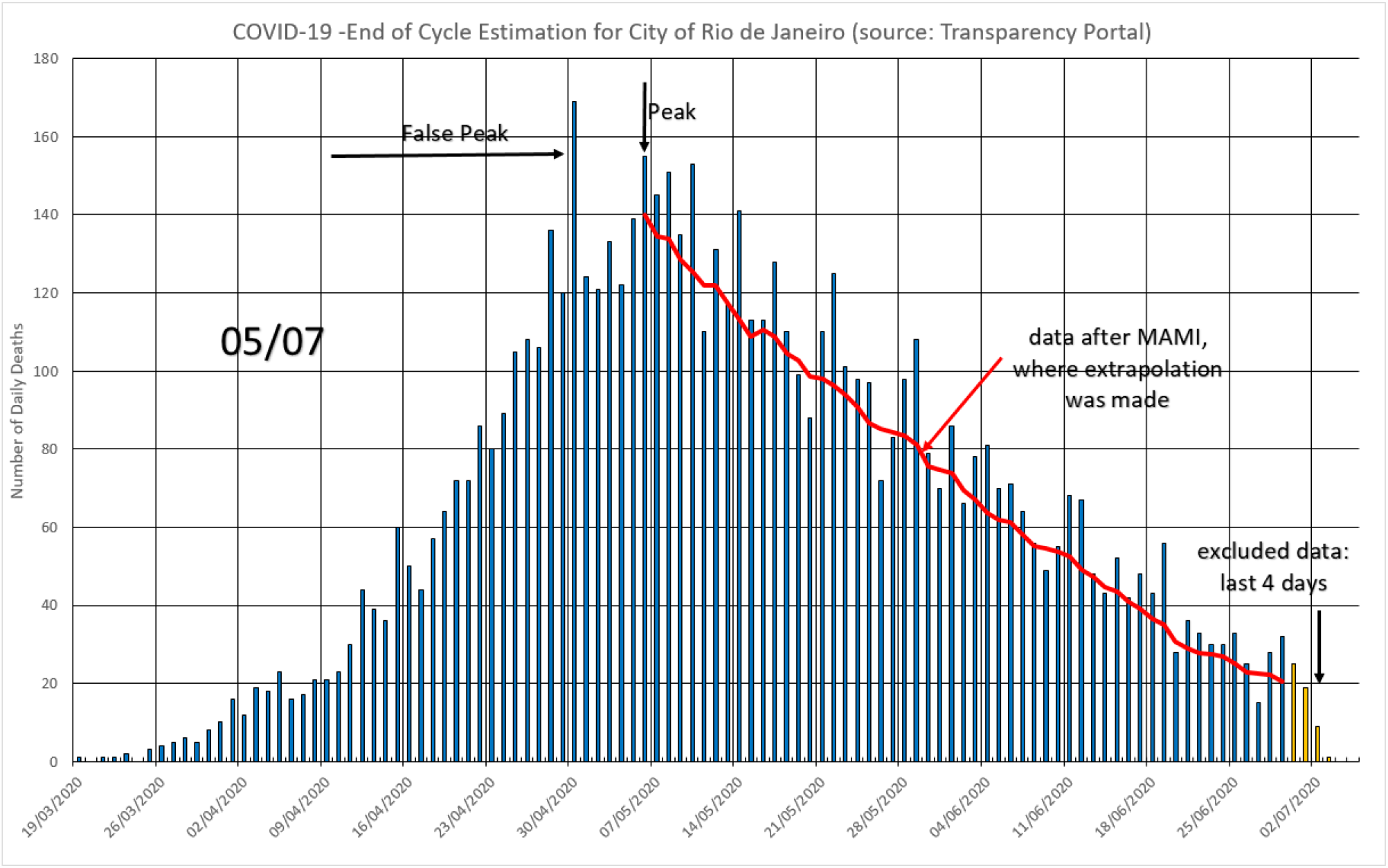
Data extrapolation for end of cycle estimation using data collected in July the 5^th^, 2020 The estimated end is at July the 07^th^.

#### 9.1.3.) Evaluation of the Rio de Janeiro’s Cycle Duration by Comparison

In item 5 of this work, several cases are compiled whose critical cycles can, at this moment, be considered as closed. They will be used for a very simple assessment, considering that there is a proportion between the time elapsed in the cycle of rise, so to speak, of the disease and that of the fall in deaths. Table 6 and 7 presents these estimates based on the cases presented. Figure 19 shows the Rio de Janeiro’s cycle, which presented twice the time of ascent as the world average (21 against 42 days). On the other hand, the decline was relatively rapid, compared to the rest of the world, although this is due to the long rise. The number of actual deaths (6918) registered at the registry office on 07/07/2020 is within the limits of Table 3, although very close to the lower limit. As for the estimated time, following the proportions listed in Table 2, the values are absurd, since the Rio de Janeiro’s cycle, for all purposes, is at the end.

**Table 6.** Cycle duration estimation for the City of Rio de Janeiro, based on Table 2 values.

| Reference Place | Multiplication Factor | Days up to the peak in Rio | Days up to the end in Rio | Predicted Day |
| --- | --- | --- | --- | --- |
| Spain | 2,1 | 42 | 88 | 02/aug/2020 |
| Germany | 3,2 |  | 134 | 17/sep/2020 |

**Table 7.** Number of deaths estimation for the City of Rio de Janeiro, based on Table 3 values.

| Reference Place | Multiplication Factor | Deaths up to the peak in Rio | Deaths up to the end in Rio | Predicted Total Deaths |
| --- | --- | --- | --- | --- |
| Sweden | 1,6 | 2588 | 4141 | 6729 |
| Suécia | 3,3 |  | 8540 | 11128 |

**Figure 19.**
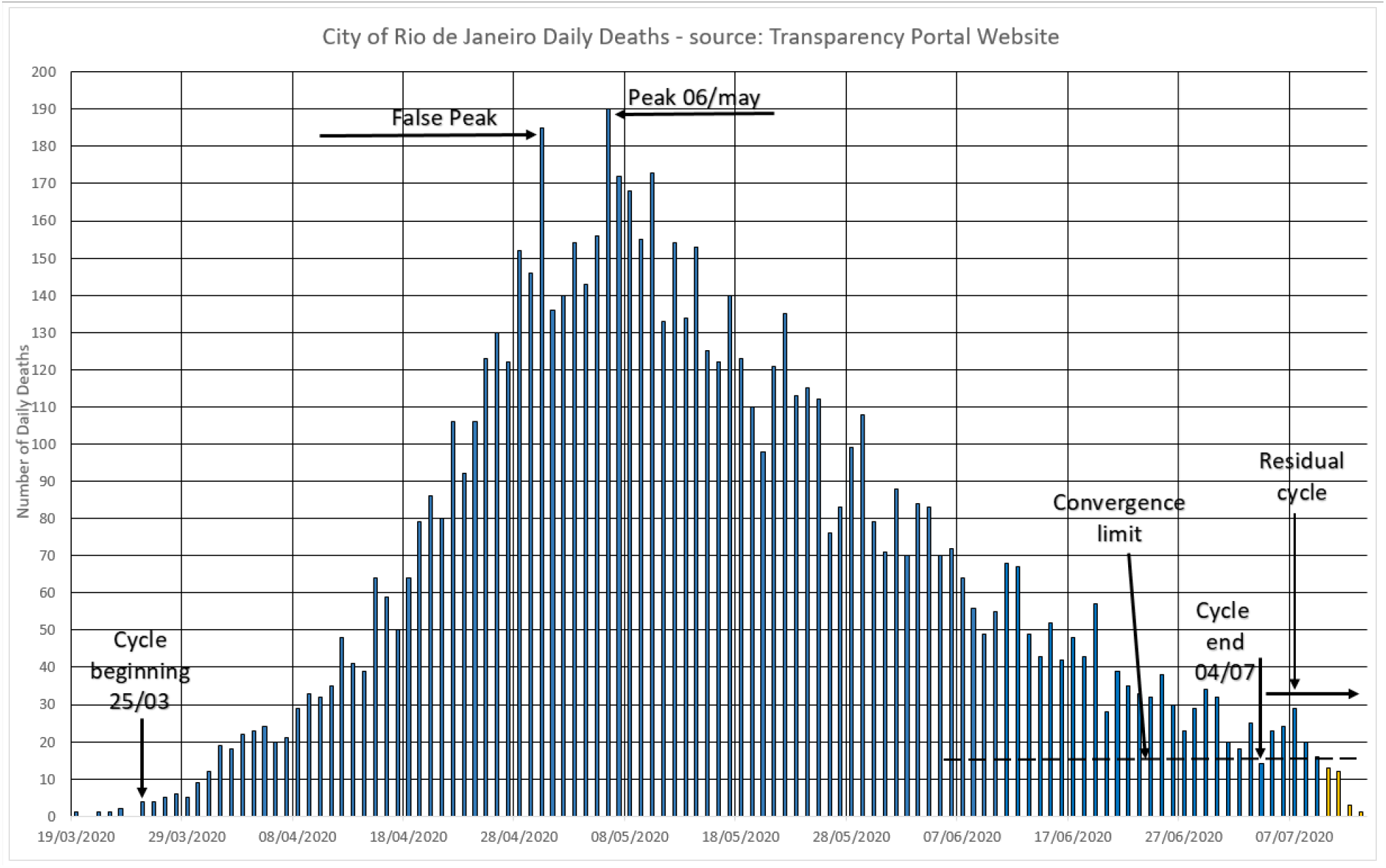
The Rio de Janeiro’s cycle, closed already by the time this work was written, presented almost symmetrical ascent and descent times and number of deaths.

## 10) Conclusions

The immediate conclusions that were obtained from the analyzes demonstrated here are listed below:

a. Given the large number of under-reporting, typical of epidemic episodes, the data considered most reliable were those related to the daily number of deaths, which were then used in this study.
b. The nature of the data currently available for studies requires preliminary numerical treatment, since most of them present the number of daily deaths that occurred on the dates on which they were recorded and not on those that actually occurred.
c. There is an observable cycle in several small and medium countries, cities and states, apparently independent of the measures taken by the local authorities. Its appearance is triangular, with the descent time that varies from 2.1 to 3.3 times longer than the ascent. And the number of deaths in the period of deceleration of the disease is 1.6 to 3.3 times greater than in the period of acceleration.
d. There are, however, among this triangular type, several cycles that are similar, once their values are made independent of the scale and dates of occurrence. Normalization allows you to use an already completed cycle to estimate the behavior of a cycle that is still evolving.
e. Brazil and other countries with vast territories and populations should not be treated as a single case, but should be studied regionally, so that the evolution of the disease cycles can be clearly understood.
f. The method using the similarity of cycles was able to estimate the end of the cycle up to 34 days before the end of the cycle, but requires that there be a similar cycle.
g. The data extrapolation method was able to estimate the end of the cycle 13 days in advance and does not depend on any data other than that offered by the Transparency Portal.
h. The comparison method was not a good cycle estimator for the city of Rio de Janeiro.The authors hope that this brief study will contribute to support the decision-making process in the current and in the next pandemics that may arise, bringing rationality to it.

## Data Availability

All the data is available in the Brazilian Ministry of Health and Brazilian National Council for Justice Transparency Portal.

https://transparencia.registrocivil.org.br/especial-covid

https://covid.saude.gov.br/

https://www.worldometers.info/coronavirus/

## Acknowledgments

The authors would like to thank Dr. Renato Nunes Teixeira, from INMETRO (National Institute of Metrology, Quality and Technology), for reviewing this work.

## APPENDIX A

### Comparison between Data from the Transparency Portal and the MoH

Both databases offer advantages and disadvantages, but both are reliable and over time will converge to similar values. To prove the veracity of this statement, the following procedure was done:

a. The daily data on deaths, released by the MoH were converted into new values by the Moving Average Method with Initial value (MAMI), that is, the average of the values of 6 days ahead, plus that of the day itself, assumes the first value in the spreadsheet and so on.
b. A curve was then drawn connecting all these points (black curve).
c. The daily data on deaths collected on the Transparency Portal website were listed and a curve linking all these points was drawn (red curve).
d. The two curves were superimposed, for comparison purposes, since they originally occur on different dates (as expected).
e. It is clear that there is a period of agreement between the two curves, which is the beginning of the epidemic, whether in Brazil or in the locality of interest. When moving to the right, the difference increases, consistent with the fact that recent records have not yet been made at the registry offices, but have already occurred at the health departments of municipalities and states, so these deaths “occurred” in the MoH but not in the notaries.

**Figure A1.**
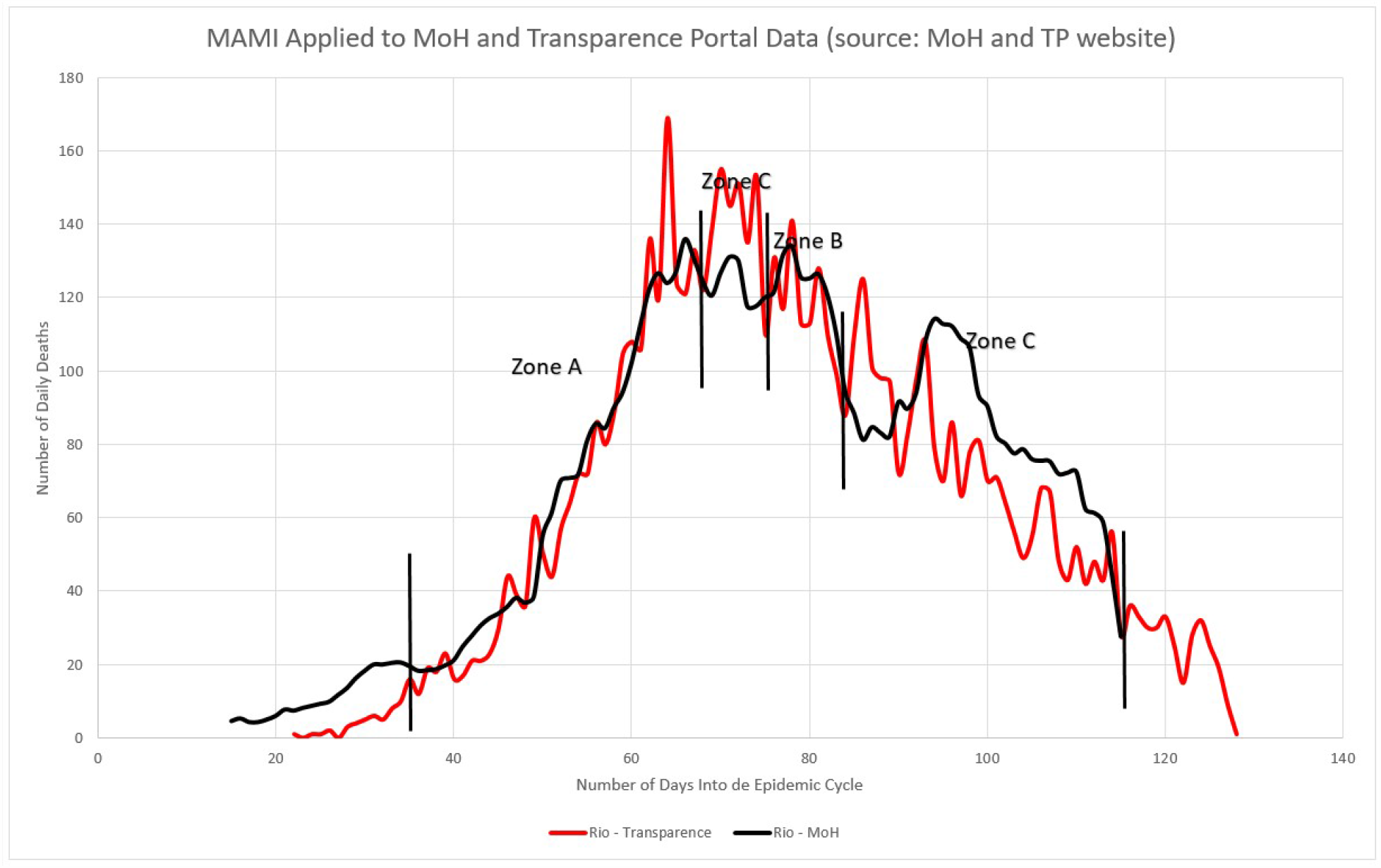
MoH (black) x TP website (red) data comparison after MAMI.

In Figure A1, Zone A can be seen, where the values of the two sources differ by an average of 7%, although they may present some value outside this range, they are negligible differences. In Zone B, this figure grows to around 9%. In Zone C, the value rises to around 15% which, although not negligible, it is still tolerable in any numerical modeling of human events. There is no equivalence between the last days, which is expected, since the two bases report past values, but with different delays. In order to perform any extrapolation using the MoH values, care must be taken to avoid using data from this final time non-concordance range.

#### A.1) The Brazilian Case

In Figure A2, Zone A can be seen, where the values of the two sources differ by an average of 1%, although they may present some value outside the unitary range, they are negligible differences. In Zone B, this figure grows to around 10%. In Zone C, the value rises to around 15% which, although not negligible, it is still tolerable in any numerical modeling of human events. In Zone D, the difference reaches about 20% thereafter, which is equivalent to the last 10 days, for Brazil, there is indeed a big difference, so to make any extrapolation, care must be taken to avoid using these values, if one uses the database of public notaries. But this phenomenon is only observable in a case as vast as that of the whole of Brazil. See the example of the city of Rio de Janeiro.

**Figure A2.**
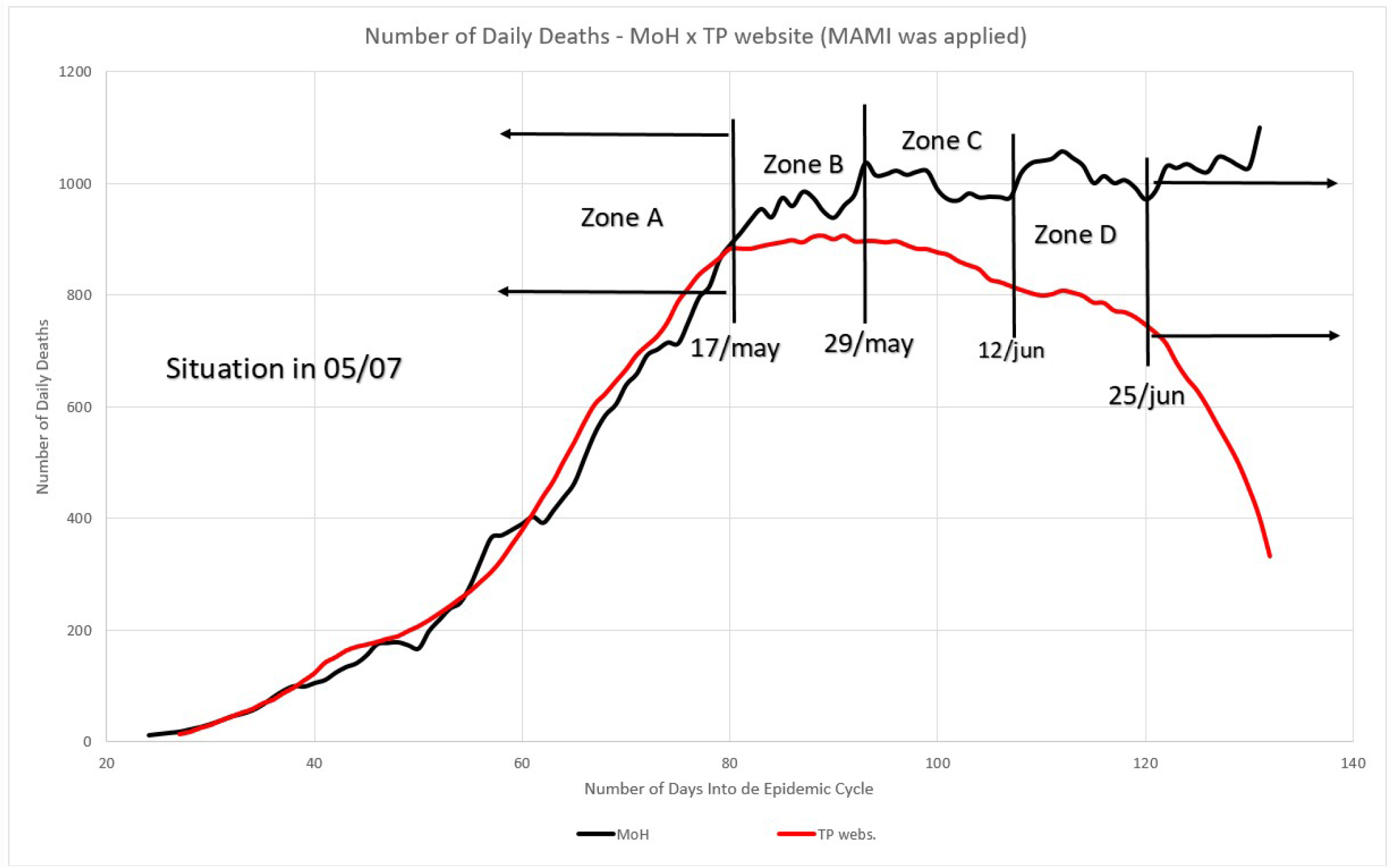
Number of daily deaths according to MoH (black line) and Transparency Portal (red).

